# Polymorphisms in Intron 1 of HLA-DRA Differentially Associate with Type 1 Diabetes and Celiac Disease and Implicate Involvement of Complement System Genes C4A and C4B

**DOI:** 10.1101/2023.06.12.23291280

**Authors:** Özkan Aydemir, Jeffrey A. Bailey, Daniel Agardh, Åke Lernmark, Janelle A. Noble, Agnes Andersson Svärd, Elizabeth P. Blankenhorn, Hemang Parikh, Anette-G. Ziegler, Jorma Toppari, Beena Akolkar, William A. Hagopian, Marian J. Rewers, John P. Mordes, TEDDY Study Group

## Abstract

Polymorphisms in genes in the human leukocyte antigen (HLA) class II region comprise the most important inherited risk factors for many autoimmune diseases including type 1 diabetes (T1D) and celiac disease (CD): both diseases are positively associated with the HLA-DR3 haplotype (*DRB1*03:01-DQA1*05:01-DQB1*02:01*). Studies of two different populations have recently documented that T1D susceptibility in HLA-DR3 homozygous individuals is stratified by a haplotype consisting of three single nucleotide polymorphisms (“tri-SNP”) in intron 1 of the *HLA-DRA* gene. In this study, we use a large cohort from the longitudinal “The Environmental Determinants of Diabetes in the Young” (TEDDY) study to further refine the tri-SNP association with T1D and with autoantibody-defined T1D endotypes. We found that the tri-SNP association is primarily in subjects whose first-appearing T1D autoantibody is to insulin. In addition, we discovered that the tri-SNP is also associated with celiac disease (CD), and that the particular tri-SNP haplotype (“101”) that is negatively associated with T1D risk is positively associated with risk for CD. The opposite effect of the tri-SNP haplotype on two DR3-associated diseases can enhance and refine current models of disease prediction based on genetic risk. Finally, we investigated possible functional differences between the individuals carrying high and low-risk tri-SNP haplotypes, and found that differences in complement system genes C4A and C4B may underlie the observed divergence in disease risk.

## INTRODUCTION

Type 1 diabetes (T1D) is an autoimmune disease that results from destruction of pancreatic islet β cells by autoreactive T cells (Wang et al., 2017). Clinical onset is preceded by a prodrome of β-cell autoimmunity, usually marked by the appearance of autoantibodies directed principally at insulin (IAA), glutamic acid decarboxylase (GADA), and islet antigen-2 (IA-2A). T1D is a non-Mendelian polygenic disorder involving the interaction of multiple gene variants, environmental factors, and immunoregulatory dysfunction (Wang et al., 2017). Only about 12% of persons with T1D have a first-degree relative (FDR) with the disease, a percentage that has remained unchanged despite increasing incidence (Dahlquist et al., 1989; Parkkola et al., 2013). The environmental factors that may precipitate disease in genetically susceptible individuals and the mechanisms by which they act have not been completely identified.

Gluten enteropathy, or celiac disease (CD), is also an autoimmune disorder. It results in atrophy of intestinal villi in the context of intestinal T cell inflammation and is associated with the presence of transglutaminase autoantibodies (tTGA) (Husby et al., 2012; Leonard et al., 2017). Although gluten is necessary for the disease it is not by itself sufficient, and CD is likely triggered by other environmental exposures (Andren Aronsson et al., 2019). Clinical disease is characterized by a range of symptoms and signs including abdominal discomfort, disordered intestinal motility, and nutrient malabsorption. Like T1D, CD is a non-Mendelian polygenic disorder involving interaction of multiple gene variants, environmental factors, and immunoregulatory dysfunction. On average, about 7.5% of persons with CD have a first-degree relative with the disorder, but the prevalence varies with the relationship, sex, and geographic location (Singh et al., 2015).

The major genetic loci for both T1D and CD susceptibility encode major histocompatibility complex (MHC) class II cell surface antigens; they are encoded by the human leukocyte antigen (HLA) complex genes on chromosome 6 (Gutierrez-Arcelus et al., 2016) Together with the T cell receptor (TCR) and an autoantigenic peptide, these glycoproteins form the “tri-molecular complex” that is central to cell-mediated immune and autoimmune responses.

T1D susceptibility is most strongly associated with polymorphisms in HLA class II genes. The major HLA class II genes that predispose to T1D are *HLA-DRB1, DQA1,* and *DQB1,* which encode the HLA DR and DQ heterodimers (Polychronakos & Li, 2011). Most T1D patients bear genetic haplotypes known as HLA-DR3 (*DRB1*03:01-DQA1*05:01-DQB1*02:01*) and HLA-DR4 (*DRB1*04:01-, *04:02-, *04:04-* or *\*04:05-DQA1*03:01-DQB1*03:02*). The DR3 haplotype can also be abbreviated as DR3-DQ2, with DR3 referring to presence of the *DRB1*03:01* gene and DQ2 referring to the heterodimeric protein product of the *DQA1*05:01* and *DQB1*02:01* genes. The DR4 haplotype represents any of the *DRB1*04*-encoded allotypes with DQ8, the heterodimeric protein product of the *HLA-DQA1*03:01* and *DQB1*03:02* genes (DR4-DQ8). T1D is also associated with HLA class I genes (Noble et al., 2010; Polychronakos & Li, 2011) and with class II *HLA-DPB1* genes (Bradfield et al., 2011). Genome wide association studies (GWAS) revealed >50 non-MHC loci associated with the disease (Bradfield et al., 2011) and more recent analyses have extended that number to >75 (Robertson et al., 2021). Because their individual contribution to total risk is small (Baranzini, 2009), the analysis of non-MHC risk loci has not improved our ability to predict disease based on genotype alone.

In CD, about 90% of patients carry DR3-DQ2, and most of the remaining CD patients carry the DR4-DQ8 haplotype (Brown et al., 2019). Fine mapping of the HLA region in persons with CD has identified additional associations that account for approximately 18% of the genetic risk (Gutierrez-Achury et al., 2015). These loci, together with the >50 known non-MHC susceptibility loci identified by GWAS, are thought to explain up to 48% of overall CD heritability (Gutierrez-Achury et al., 2015).

The DQ2 and DQ8 haplotypes positively associated with CD are carried on the DR3-DQ2 and DR4-DQ8 haplotypes, respectively, that are predisposing for T1D. The occasional presence of both T1D and CD in the same individual is therefore not surprising. The rate at which CD occurs in persons with T1D varies substantially among various populations and reportedly ranges from 2.5% to 16.4% (De Vitis et al., 1996; Szaflarska-Poplawska, 2014). In the HLA-selected TEDDY study T1D autoimmunity usually preceded CD autoimmunity in cases that had both (W. Hagopian et al., 2017).

In the case of T1D, HLA genotyping can identify persons at risk for the disease, but only about 1 in 15 (∼7%) individuals with even the highest-risk HLA genotype identified in Europeans (HLA-DR3/DR4 heterozygotes) develop the disorder (Rewers et al., 1996). Similarly, only about 3% of persons with DQ2 or DQ8 develop celiac disease (Brown et al., 2019). Recent studies that coupled genetic analysis with phenotypic markers have improved our ability to predict islet autoantibodies and T1D in the general population (Bonifacio et al., 2018), but additional genetic risk remains to be discovered (Pierce et al., 2013). Understanding of the detailed genetic risk conferred by HLA-DR and -DQ-encoding genes remains incomplete and provided the impetus for these studies.

We have previously reported that susceptibility to T1D among persons homozygous for the high-risk DR3-DQ2 haplotype is powerfully modulated by variation within intron-1 of the *HLA-DRA* gene (Aydemir et al., 2019), a gene that, unlike most HLA-encoding genes, is considered essentially invariant and largely ignored in HLA association studies. This variation comprises three single nucleotide polymorphisms (SNPs): rs3135394, rs9268645, and rs3129877 (referred to here as the “tri-SNP”) in a 100 bp interval within intron 1. We called the T1D risk haplotype (nucleotides AGG) “010” using (1) to designate the reference and (0) alternate alleles, respectively. T1D protection was conferred by the “101” haplotype (GCA). The effect of the tri-SNP on T1D susceptibility was discovered in an analysis of samples drawn from the Type 1 Diabetes Genetics Consortium (T1DGC), the Swedish Better Diabetes Diagnosis (BDD) study, and the Swedish Diabetes Prediction in Skåne (DiPiS) study (Aydemir et al., 2019). The relative T1D risk of the 010/010 genotype, compared to the homozygous 101/101 genotype, was substantial in both the T1DGC and Swedish DR3/3 cohorts (odds ratio 4.65, p=1.69 × 10^−13^). This association was later independently replicated in a separate cohort of DR3-DQ2/DR3-DQ2 individuals from Finland (Nygard et al., 2021). These investigators reported that the 010/010 (AGG/AGG) or 010/x (AGG/x) tri-SNP haplotypes were significantly more common in patients with T1D (OR = 1.70, CI 95%=1.15–2.51, p = 0.007) than in non-diabetic controls.

Here we report an expanded analysis of the effect of the tri-SNP on the appearance of islet autoimmunity and the subsequent progression to T1D using data from the large population enrolled in The Environmental Determinants of Diabetes in the Young (TEDDY) study (Teddy Study Group, 2008). In addition, because CD is also documented in the TEDDY study, the novel and unexpected discovery was made that the tri-SNP 101 genotype that confers T1D protection was associated with increased susceptibility to CD.

## METHODS

### Study Population

Data were obtained from participants in the TEDDY prospective cohort study whose primary goal is to identify environmental causes of T1D. It includes three clinical research centers in the United States (Colorado, Georgia/Florida, Washington) and three in Europe (Finland, Germany, and Sweden), and has been described in detail (Teddy Study Group, 2007, 2008). Briefly, participation in the study was offered to the parents of all newborns at each research center. Inclusion criteria were based on HLA genotype and are described below. Separate written informed consents were obtained for all study participants from a parent or primary caretaker for genetic screening and for participation in prospective follow-up. The study was approved by local Institutional Review Boards and is monitored by an external advisory board formed by the National Institutes of Health.

#### Inclusion Criteria and HLA Typing

Genotype screening (W. A. Hagopian et al., 2011) was conducted using either a dried blood spot punch or a small volume whole blood lysate specimen as described (Dantonio et al., 2010). Infants were eligible for the study if they had 2 high-risk HLA genotypes or had a first degree relative with T1D and at least one high-risk HLA genotype. Inclusion criteria and the HLA nomenclature used is detailed in the Supplementary Methods. Screening blood samples were generally obtained at birth from cord blood. Other potential participants, especially first-degree relatives of T1D participants at the Washington site, were screened using heel stick capillary samples up to the age of 4 months. This exception was made to maximize the number of newborn relatives participating in this study. After polymerase chain reaction (PCR) amplification of exon 2 of the HLA Class II gene (DRB1, DQA1 or DQB1), alleles are identified either by direct sequencing, oligonucleotide probe hybridization, or other genotyping techniques as described (W. A. Hagopian et al., 2011). Additional typing to sufficiently identify certain DR-DQ haplotypes was specific to each clinical center.

When a TEDDY participant was 9–12 months of age, HLA status was confirmed by genotyping at increased resolution of HLA-DRB1, DQA1, and DQB1 at the central HLA reference laboratory at Roche Molecular Systems, Oakland, CA (Erlich et al., 1991). SNPs from the Illumina Immuno BeadChip (manifest file: Immuno_BeadChip_11419691.bpm from Illumina, San Diego, CA, USA) were also assessed by the Center for Public Health Genomics at University of Virginia as described previously (Krischer et al., 2017).

### Study Design

#### Primary Outcome Definition

The primary TEDDY outcome variable was development of persistent, confirmed islet autoimmunity (IA). IA was assessed every 3 months through four years of age. Persistent autoimmunity was defined by the confirmed (two reference labs in agreement) presence of any one of three islet autoantibodies: autoantibody to insulin (IAA), glutamic acid decarboxylase (GADA), or insulinoma antigen-2 (IA-2A) on two or more consecutive visits. Date of persistent autoimmunity was defined as the draw date of the first sample of the two consecutive autoantibody-positive samples.

#### Islet Autoantibodies

Islet autoantibodies were the first primary end-point in the TEDDY study. Islet autoantibodies to insulin, GAD65, and IA-2 were measured in two laboratories by radiobinding assays (Babaya et al., 2009; Bonifacio et al., 2010). In the U.S., all sera were assayed at the Barbara Davis Center for Childhood Diabetes at the University of Colorado Denver; in Europe, all sera were assayed at the University of Bristol, the U.K. Assays in both laboratories have previously demonstrated high sensitivity, specificity and concordance (Torn et al., 2008). All positive islet autoantibodies and 5% of negative samples were re-tested in the other reference laboratory and deemed confirmed if concordant. To optimize concordance, harmonized assays for GADA and IA-2A replaced earlier assays in January, 2010.^1^ To distinguish maternal antibodies from IA in the child, the IA status of the mother was measured when the child was aged 9 months, and the child’s IA status was measured at 3 months of age and then every three months until 18 months of age. If the child had positive IA during this time, the status was listed as ‘pending’ until 18 months of age, at which time the child’s IA status was determined based on both maternal and child IA over the first 18 months of the study. If maternal antibodies were present, the child was not considered persistently IA positive until positive at/after 18 months unless the child had a negative sample prior to their first positive sample. A recognized limitation of this approach is that true IA positivity during the first 18 months of life that waned could have been masked by maternal antibodies.

#### Diagnosis of T1D and CD

T1D and CD were the second primary end-points in the TEDDY study. At the time of clinical diagnosis of diabetes, data were collected outside the TEDDY clinics (care providers or clinics) using a standardized case report form requiring documentation to fulfill American Diabetes Association criteria for classification of T1D (Elding Larsson et al., 2014).

TEDDY children were screened for tTGA using radiobinding assays from 2 years of age and annually thereafter. TEDDY samples from the US sites were analyzed at the Barbara Davis Center and samples from the European sites at the Bristol University, which was chosen as the reference laboratory as previously described (Liu et al., 2014). All samples from US children with a tTGA level >0.01 units were sent to the Bristol laboratory for analysis. Children with a positive tTGA had additional samples collected at three-month intervals to 48 months of age, and at 6 month intervals after the 48-month visit. In addition, all tTGA positive children had previously collected samples analyzed to find the closest time-point to conversion. Children with a positive tTGA result in two consecutive samples were defined as having celiac disease autoimmunity (CDA) (i.e., primary outcome) and referred to local health care providers for follow-up and clinical evaluation of CD. The decision to perform a diagnostic intestinal biopsy was outside the TEDDY protocol, but it was recommended by the TEDDY investigators of Celiac Disease Committee to biopsy CDA children with tTGA levels 30 units or greater in CDA children with gastrointestinal symptoms regardless of tTGA level. Diagnosis of CD per se was defined as an intestinal biopsy showing a Marsh score >1, or if a biopsy was not performed, having a mean tTGA level of 100 units or greater in two consecutive samples (i.e., secondary outcome), respectively.

### Statistical and Computational Analyses

#### Whole Genome Sequencing

Whole genome sequencing (WGS) was conducted on the subjects based on the IA and T1D nested case–control (NCC) studies with targeted 30× coverage by Macrogen, Inc, Rockville, MD, U.S.A as described previously (Törn et al., 2022). Joint variant discovery and genotype calling was performed by combining TEDDY and TOPMed WGS data using topmed_variant_calling pipeline (https://github.com/statgen/topmed_variant_calling, (Taliun et al., 2021))

#### Determination of tri-SNP haplotype

Tri-SNP haplotypes were extracted from the ImmunoChip or WGS phased variant call file using scikit-allel software v1.3.5 (https://scikit-allel.readthedocs.io/en/stable/).

#### Determination of B8-DR3 haplotype

Extended haplotype status of the individuals was imputed based on a subset of 203 SNPs whose non-reference alleles were associated with the B8-DR3 haplotype (*A*01:01-B*08:01-DRB1*03:01*) (Gourraud et al., 2014). The ImmunoChip variant calls contained 79 of these variants whereas the WGS data contained 200. The individuals were assigned a B8-DR3 dosage based on the mode of the alternative allele counts of all SNPs, *i.e*. 0 if most SNPs were called homozygous reference genotype, 1 if most loci were heterozygous and 2 if most loci were homozygous alternate allele.

#### Genetic ancestry inference

Out of 7759 individuals 5277 (68.0%) had self-reported race/ethnicity information as African American (65), Hispanic (558) and White non-Hispanic (4654). These were assigned to AFR, AMR and EUR superpopulation groups, respectively. The remaining 2482 individuals’ population membership was inferred using the software Kinship-based INference for Gwas (KING v2.2.7, (Manichaikul et al., 2010)) ancestry inference commands (`king -b reference.bed,samples.bed --pca --projection`). Briefly, the program uses 2409 samples from the 1000 genomes project with known population information to create Principal Components (PCs) and projects the study samples onto these PCs based on their genetic variation data (from ImmunoChip and Whole Genome Sequencing for TEDDY samples). Python sklearn library’s (v0.24.2) RandomForestClassifier module was used to assign population membership using the PCs generated by KING. The classifier’s performance was assessed by splitting the 2409 individuals from 1000 genomes project with known ancestry into training (70%) and test (30%) sets which showed an accuracy of 99.6%. For ancestry assignment of study samples, all 2409 samples from 1000 Genomes Project were used as the training data and all 7903 TEDDY samples (all samples with genetic data including those that were not in the final data set. e.g. with ineligible HLA types) as test data. Study samples where the ancestry information was reported were compared to the classifier predictions which showed 95.6% concordance. The inferred population information was used only for the 2482 samples that did not have the self-reported ancestry data.

#### Statistical model

The Cox Proportional Hazards (PH) method was used to analyze the impact of tri-SNP haplotypes on primary and secondary T1D and CD disease outcomes. T1D diagnosis, islet autoimmunity (IA), first-appearing islet autoantibody, celiac diagnosis (CD) and celiac disease autoantibody (CDA) status were defined as events in separate models; age of the subject (in months) was used as the time component. When the event was not observed, age at the latest clinic visit (for CD or T1D) or at the last negative serum sample collection (for IA and CDA) was used as the right-censor time. In the first appearing autoantibody models, samples were right-censored if any other antibody than the one in the model outcome first appeared.

Known risk factors including HLA type, sex, having a first degree relative with the disease and genetic variants previously shown to be associated with each outcome (Krischer et al., 2017; Sharma et al., 2016, 2018), as well as potential confounders including ancestry/ethnicity and the country of residence were included in the model to adjust for them. Tri-SNP 101 allele was modeled as a numerical variable (0, 1 or 2 alleles) representing an additive genetic effect, and hence, its hazard ratio values should be interpreted as HR per each additional allele. Similarly, the known GWAS loci were encoded numerically (0, 1, 2 alternate alleles). For the categorical covariates HLA type, sex, country and genetic ancestry the baselines were assigned to DR4/DR8, female, USA and EUR, respectively. Cox PH modeling was carried out using the Lifelines Python library v0.26.3 (Davidson-Pilon, 2019).

#### RNA Sequence Analysis

The RNA samples were prepared using Illumina’s TruSeq Stranded mRNA Sample Prep Kit from the whole blood samples. RNA sequencing was conducted using the Illumina HiSeq4000 platform with paired-end 2 x 101 bp reads with a targeted 50 million reads per sample by the Broad Institute, Cambridge, MA. Gene expression was quantified using salmon software v1.6.0 (Patro et al., 2017) and Gencode v39 transcript set. Differential gene expression analysis was carried out using limma v3.48.3 (Ritchie et al., 2015) with the design formula “∼ batch + age + sex + triSNP 101” to account for differences due to sequencing batch, age and the sex of each sample. Age was modeled as a categorical: younger or older than 1 year old as the major difference due to age was in children younger than 1 based on PCA analysis of gene expression data (not shown). Multiple samples from the same individual were accounted for by using the individual as the blocking factor as recommended in limma user manual. Independent effect of triSNP-101 was extracted from the model using the *topTable* function and coefficient triSNP 101.

#### Copy Number Analysis

A bed file containing 1 kilobase non-overlapping windows for MHC region (chromosome 6 positions 28510000-33482000) was created. MHC locus read coverage for each sample was extracted from WGS reads mapped to human genome assembly hg38 using samtools v1.16.1 bedcov command and the MHC bed file. “-Q 30” option was used for counting uniquely mapping reads. The average normalized coverage for C4A and C4B genes using uniquely mapping reads (Figure 3), as well as combined C4 average coverage using all reads (Supplementary Figure 13A) were calculated. We estimated overall C4 gene copy number (GCN) based on total reads mapping to the C4 region containing C4A and C4B (Supplementary Figure 13B, Supplementary Methods). The relative GCNs of C4A and C4B were determined based on the ratio of reads that uniquely mapped to either C4A or C4B (Supplementary Figure 13C, Supplementary Methods).

## RESULTS

### Study Population Demographics

The TEDDY database analyzed here was frozen as of 31 October 2021 and the present analysis represents a median follow up of 12.7 years (IQR=10.9-14.4). 382 (5.0%) children had been diagnosed with T1D and 617 (8.0%) with CD. The TEDDY study is ongoing and about 75% of the research subjects have still to age out (15 years of age) of the study.

There were 7759 study subjects with HLA and tri-SNP typing data available. Ancestry distribution was 89.6% European (EUR, N=6953), 9.1% Ad Mixed American (AMR, N=707), 1% African (AFR, N=82), 0.2% South Asian (SAS, N=14) and <0.1% East Asian (EAS, N=3).

The distribution of tri-SNP haplotypes and HLA genotypes is shown in Table 1. As anticipated, given the study inclusion criteria, most (N=7503) participants were homozygous for either DR3 or DR4, heterozygous for DR3/DR4, or heterozygous for DR4/DR8. With respect to the distribution of tri-SNP haplotypes in this population, nearly all subjects had at least one 101 or 010 tri-SNP; only 22 (0.3%) did not. The 101 tri-SNP haplotype was observed almost exclusively coupled to the DR3-DQ2 haplotype; 99.6% (2357/2366) of the chromosomes from 101/101 homozygous individuals carried the DR3-DQ2 haplotype. Similarly, 101 is highly enriched in the DR3 population; 84% (2716/3232) of the chromosomes from DR3 homozygous individuals had 101, in line with our previous report (Aydemir et al., 2019). Approximately 99% of the DR3 homozygous population carried either 010 or 101 (3203/3232). We also observed that 98.2% of the DR4/DR4 homozygous population carried 010 (3007/3062).

**Table 1.** Distribution of HLA and Tri-SNP haplotypes among TEDDY samples.

|  | Tri-SNP Genotype |  |  |  |  |  |  |  |  |  |  |  |
| --- | --- | --- | --- | --- | --- | --- | --- | --- | --- | --- | --- | --- |
|  | 010/010 | 010/101 | 101/101 | 000/010 | 001/010 | 000/101 | 000/000 | 000/001 | 001/101 | 011/101 | 010/011 | All |
| HLA |  |  |  |  |  |  |  |  |  |  |  |  |
| DR3/DR3 | 62 | 348 | 1177 | 13 | 2 | 11 | 0 | 0 | 2 | 1 | 0 | 1616 (20.8%) |
| DR3/DR4 | 408 | 2550 | 3 | 19 | 8 | 30 | 0 | 1 | 1 | 1 | 1 | 3022 (38.9%) |
| DR4/DR4 | 1480 | 5 | 3 | 39 | 3 | 1 | 0 | 0 | 0 | 0 | 0 | 1531 (19.7%) |
| DR4/DR8 | 14 | 0 | 0 | 1297 | 5 | 0 | 17 | 1 | 0 | 0 | 0 | 1334 (17.2%) |
| DR1/DR4 | 0 | 0 | 0 | 15 | 143 | 0 | 0 | 2 | 0 | 0 | 0 | 160 (2.1%) |
| DR4/DR13 | 0 | 0 | 0 | 0 | 56 | 0 | 0 | 1 | 0 | 0 | 0 | 57 (0.7%) |
| DR3/DR9 | 2 | 16 | 0 | 0 | 0 | 0 | 0 | 0 | 0 | 0 | 0 | 18 (0.2%) |
| DR4/DR9 | 13 | 1 | 0 | 0 | 0 | 0 | 0 | 0 | 0 | 0 | 0 | 14 (0.2%) |
| DR4/DR4*030X/020X | 4 | 0 | 0 | 0 | 0 | 0 | 0 | 0 | 0 | 0 | 0 | 4 (0.1%) |
| DR4/DR4*030X/0304 | 3 | 0 | 0 | 0 | 0 | 0 | 0 | 0 | 0 | 0 | 0 | 3 (0.0%) |
| All | 1986<br>(25.6%) | 2920<br>(37.6%) | 1183<br>(15.2%) | 1383<br>(17.8%) | 217<br>(2.8%) | 42<br>(0.5%) | 17<br>(0.2%) | 5<br>(0.1%) | 3<br>(0.0%) | 2<br>(0.0%) | 1<br>(0.0%) | 7759 (100%) |

Cox PH analysis requires each level of the categorical variables to have at least one event and one non-event data point. We removed the samples that belonged to categories with too few individuals (<20) to accommodate this requirement. These included samples from: HLA classes DR3/DR9 (n=18), DR4/DR9 (n=14), DR4/DR4*030X/020X (n=4), DR4/DR4*030X/0304 (n=3); populations SAS (n=14) and EAS (n=4). 7703 samples remained in the final set. Descriptive characteristics of the final data set with respect to each outcome studied are provided in Supplementary Tables 1-6.

### Tri-SNP association with T1D diagnosis

5.2% (397/7703) of the children were diagnosed with T1D. 65 were excluded from the time-to-T1D analysis due to missing data for controlled GWAS associations, leaving the final data set for this analysis at 7638, of which 5.2% (395) were diagnosed with T1D. Our model indicated that tri-SNP 101 reduced the risk of developing T1D (HR=0.54, CI=0.41-0.72, p=1.52e-5, Figure 1A). Having an affected first degree relative (FDR) and HLA DR3/DR4 genotype were the largest risk factors, as expected, and other controlled covariates were confirmatory of previous reports (Krischer et al., 2017; Sharma et al., 2018) (Supplementary Figure 1, Supplementary Data, Supplementary Data Table 1).

**Figure 1.**
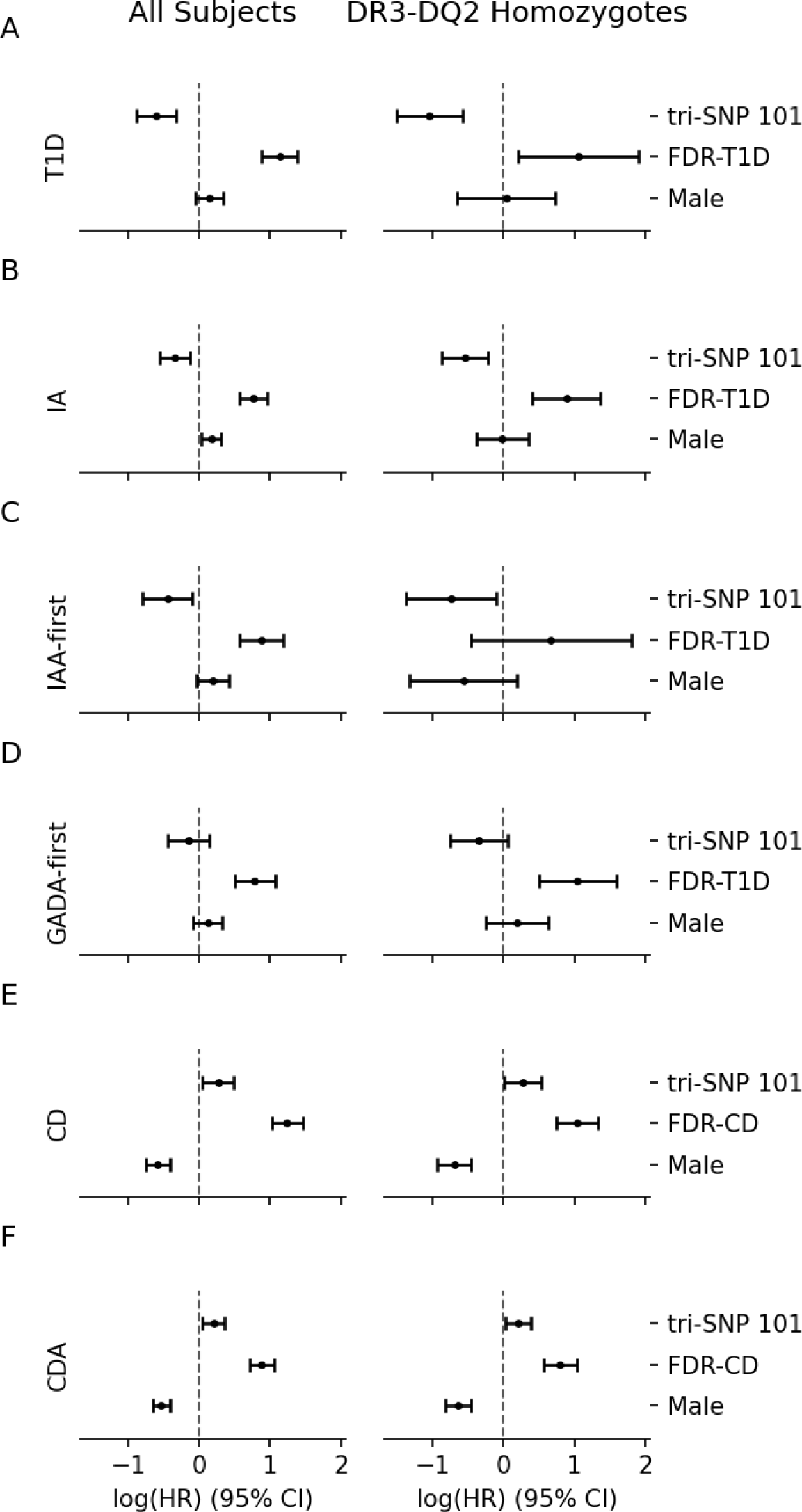
Cox PH regression results for all tested outcomes. Log hazard ratios (log(HR)) and 95% confidence intervals of the tri-SNP 101 haplotype as well as the known risk factors FDR and sex from the Cox PH models for outcomes T1D (A), IA (B), IAA-first (C), GADA-first (D), CD (E) and CDA (F), using the entire cohort (left) or only the DR3-DQ2 homozygote individuals (right). Dashed vertical line at 0 indicating an HR of 1 (log(HR) = 0), i.e. no effect on risk. Left side of the vertical line indicates reduced risk vs increased risk on the right side. Whiskers indicate 95% CI around HR. The model assesses the independent risk/protection afforded by each covariate compared to the baseline for categorical covariates FDR and sex for which the baselines are having no FDR and female sex, respectively. Tri-SNP 101 is modeled numerically, so the HR reported is per each additional 101 allele.

Given the strong linkage between tri-SNP 101 and HLA DR3 haplotypes (Table1), we performed a separate Cox PH regression using only DR3 homozygous samples for this and each subsequent study outcome. When the analysis was restricted to DR3-DQ2 homozygous children (Supplementary Figure 2, Supplementary Data Table 2), the protective effect of the 101 tri-SNP was even stronger (HR=0.35, CI=0.22-0.56, p=1.06e-5, (Supplementary Figure 2, Supplementary Data, Supplementary Data Table 2), confirming our initial report (Aydemir et al., 2019) and the confirmatory Finnish report (Nygard et al., 2021).

### Islet Autoantibody (IA) Development

At least one IA (the primary TEDDY outcome variable) was detected in 858/7703 (11.1%) children from the dataset. 89 children had missing data for controlled GWAS genotypes and were not included in the Cox PH model for IA outcome (Supplementary Figure 3, Supplementary Data Table 3). Our model showed a significantly reduced independent risk associated with the tri-SNP 101 haplotype (HR=0.71,CI=0.58-0.87, p=0.001), each protective allele conferring 29% reduction in risk of developing islet autoantibodies (Figure 1B). Among the controlled covariates, having an FDR with T1D, HLA DR3/DR4 genotype were significant risk factors for IA; and male subjects were found to be at marginally higher risk of developing IA (Supplementary Figure 3, Supplementary Data, Supplementary Data Table 3), consistent with previous TEDDY reports (Krischer et al., 2017; Sharma et al., 2018). Other controlled covariates were also in line with the previous reports (Supplementary Figure 3, Supplementary Data, Supplementary Data Table 3).

When analysis was restricted to DR3 homozygous individuals the protective effect of the tri-SNP 101 haplotype against development of IA was stronger (HR=0.58, CI=0.42-0.80, p=0.001), each allele conferring 42% reduction in risk. Having an FDR with T1D remained a significant additional risk factor within this group, but other controlled covariates were no longer significant (Supplementary Figure 4, Supplementary Data, Supplementary Data Table 4).

### First Appearing Islet Autoantibody

We assessed the effect of tri-SNP 101 on two T1D autoimmunity endotypes, based on the first appearing islet autoantibody being insulin autoantibody (IAA) or glutamic acid decarboxylase autoantibody (GADA) in separate models, controlling for the same covariates as in the IA analysis. Although tri-SNP-101 trended towards protection against GADA-first outcome, this effect did not reach statistical significance either in the entire dataset (HR=0.87, CI=0.64-1.17, p=0.34, Figure 1D, Supplementary Data Table 5) or in DR3-DQ2 homozygotes (HR=0.71, CI=0.47-1.07, p=0.10, Figure 1D, Supplementary Data, Supplementary Data Table 6). IAA-first risk was significantly reduced by tri-SNP 101 both in the entire data set (HR=0.64, CI=0.45-0.91, p=0.01) and the DR3-DQ2 homozygotes (HR=0.48, CI=0.25-0.91, p=0.03, Figure 1C, Supplementary Figures 7, 8, Supplementary Data, Supplementary Data Table). We note that since the incidence of IAA-first among DR3-DQ2 homozygotes at 1.8% (29/1578) is relatively low, samples from African genetic background (N=32, incidence=0), and one low frequency GWAS locus (Supplementary Data) were excluded from this analysis to accommodate the model requirements.

### Celiac Diagnosis (CD)

Among the study subjects 617 (8.0%) were diagnosed with CD. 24 children had both T1D and CD which corresponded to 3.9% of all CD samples and 6.0% of all T1D samples. 6530 children had complete information on all covariates for CD and these were used in the Cox PH time-to-CD analysis (Supplementary Figure 9, Supplementary Data Table 9).

In contrast to its protective effect against T1D, tri-SNP 101 was a significant risk factor (HR=1.32, CI =1.06-1.64, p=0.01, Figure 1E) for CD. As previously reported, female sex, FDR, gluten intake and HLA-DR3 and HLA-DR4 haplotypes were significant risk factors (Andren Aronsson et al., 2019), (Figure 1E, Supplementary Figure 9). Previously identified GWAS associations (Sharma et al., 2016) were also observed in our analysis (Supplementary Figure 9). Individuals assigned to AMR genetic ancestry were at lower risk of CD (HR=0.62, p=0.04) compared to the baseline EUR population.

The increased CD risk associated with the 101 tri-SNP haplotype remained significant (HR=1.32, CI=1.02-1.70, p=0.04) when the analysis was restricted to HLA-DR3 homozygotes (Figure 1 E, Supplementary Figure 10, Supplementary Data Table 10).

### Celiac Disease Autoimmunity (CDA) Development

6709 children had CDA data available. 19.2% (1294) were positive for CDA. 174 showed both CDA and IA corresponding to 13.4% of all CDA and 21.3% of all IA individuals. 6557 children had complete information for all covariates used in the statistical model (Supplementary Figure 11, Supplementary Data Table 11).

Similar to its effect on CD and contrary to T1D and IA, the tri-SNP 101 haplotype was a significant risk factor (HR=1.23, CI=1.06-1.44, p=0.008) for CDA (Figure 1F). Other risk factors included FDR with celiac disease, female sex, HLA-DR3 and DR4 haplotypes, gluten intake, European genetic ancestry, all of which have been reported previously (Andren Aronsson et al., 2019; Singh et al., 2015) (Figure 1F, Supplementary Figure 11).

Tri-SNP 101 remained a risk factor when the analysis was restricted to DR3-DQ2 homozygotes (HR=1.23, CI=1.02-1.48, p=0.03, Supplementary Figure 12, Supplementary Data Table 12).

### Comparison to HLA-DR3 B8 extended haplotype

It has been reported that two conserved extended haplotypes carrying DR3 (B8-DR3) and B18-DR3 differentially contribute to the risk for T1D and CD in the Basque population (Bilbao et al., 2006). Our analysis shows that the tri-SNP 101 haplotype is weakly correlated with the CD-predisposing B8-DR3 extended haplotype (R^2^=0.32 for DR3 homozygotes, data not shown). However, when the Cox proportional hazard model was fitted using the B8 dosage instead of the tri-SNP, the associations with T1D (B8 HR=0.64 vs tri-SNP HR=0.54, Supplementary Data Table 13) and IA (B8 HR=0.80 vs tri-SNP HR=0.71, Supplementary Data Table 14) were weaker, and no significant association was detected either for CD (HR=1.11, p=0.12, Supplementary Data Table 15) or for CDA (HR=1.07, p=0.20, Supplementary Data Table 16).

### Differential Gene Expression

We explored the leukocyte gene expression differences within 538 samples from 129 DR3-DQ2 homozygous individuals of our study cohort. Expression of 68 genes were found to be significantly affected by the tri-SNP 101 haplotype (adjusted-p < 0.01, Supplementary Data Table 17). 8 of 10 of the most significant changes were affected genes located in the MHC loci.

The expression of complement system genes C4A and C4B was inversely affected by the tri-SNP 101 dosage (Figure 2). Whereas C4A gene expression decreased (p=1.46E-22), C4B significantly increased (p=5.32E-13) with increasing tri-SNP 101 dosage.

**Figure 2.**
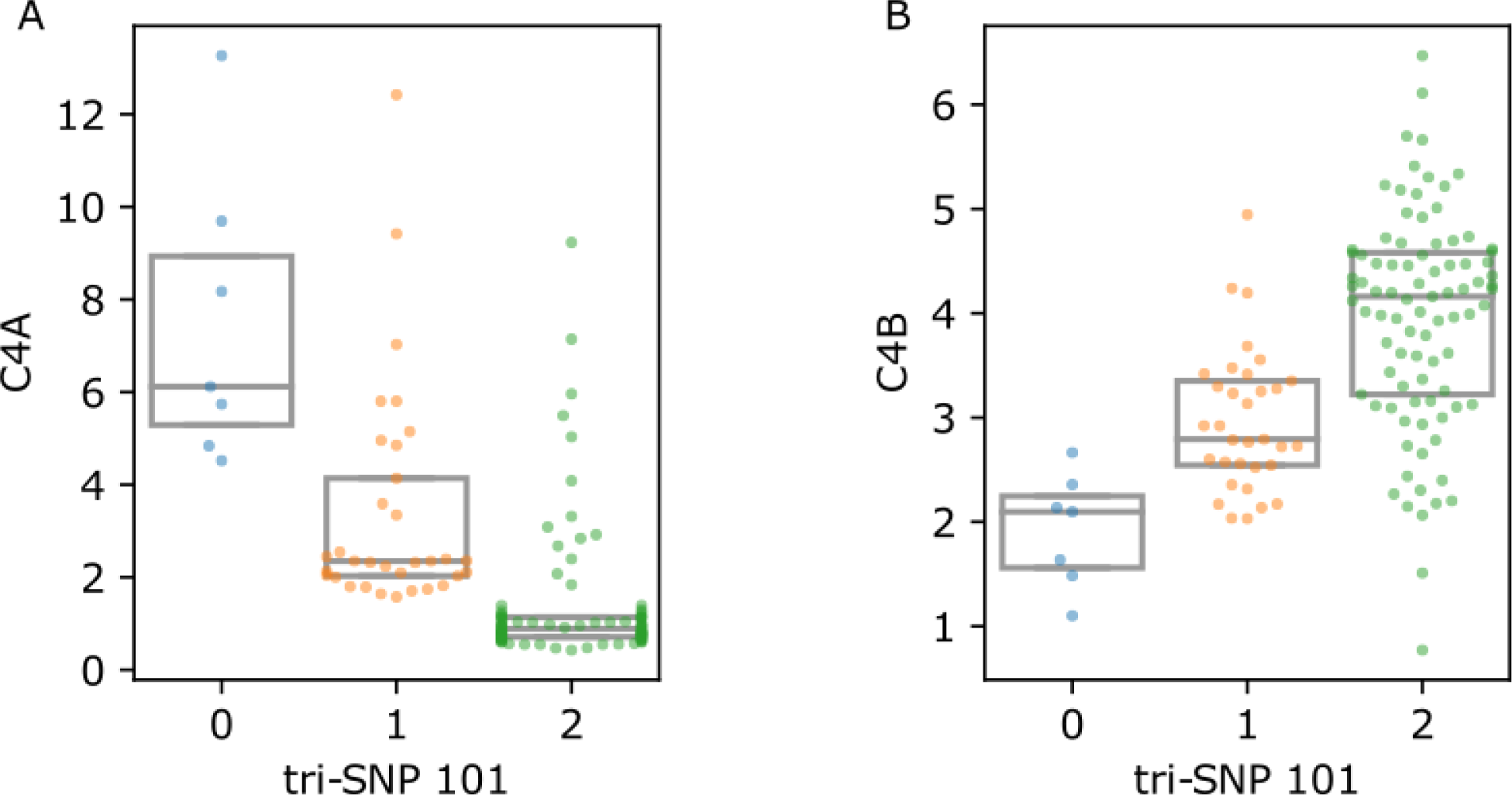
C4 gene expression values with respect to tri-SNP. Count per million (CPM) values in 129 DR3 homozygous individuals showing decreasing C4A and increasing C4B gene expression as tri-SNP 101 allele count increases. Each point represents the median CPM value of multiple samples from one individual. Boxes represent the interquartile range (IQR) and midlines mark the median value.

### Copy Number Variation in C4 Locus

The C4 locus has been reported to have frequent copy number variation and associations with autoimmunity (Li et al., 2017). We analyzed the whole genome sequencing data of 188 DR3-DQ2 homozygous individuals from our cohort to investigate whether there are copy number changes in C4A or C4B. Read coverage data showed reduced coverage of C4 genes in some samples indicating presence of gene deletions (Supplementary Figure 13A). Due to extensive sequence identity between C4A and C4B, determination of C4A and C4B copy numbers was based only on reads mapping uniquely to either gene (Figure 3). We identified frequent C4A deletions: 56.9% (107/188) of the samples were C4A null and 23.4 % had only a single C4A (Supplementary Table 8). All 107 C4A null samples were homozygous for tri-SNP 101 genotype indicating a strong association (chi-squared p-value = 5.59E-38). C4B deletions were also common, although not as frequent as C4A. There was only one C4B null sample and 22.8% (43/188) of the samples had a single C4B gene (Supplementary Table 9). We observed a positive association between tri-SNP 101 and C4B copy number (chi-squared p-value = 1.89E-20) in contrast to the negative correlation with C4A. We also noted that 89.4 % (168/188) of samples had total C4 gene copy number <4.

**Figure 3.**
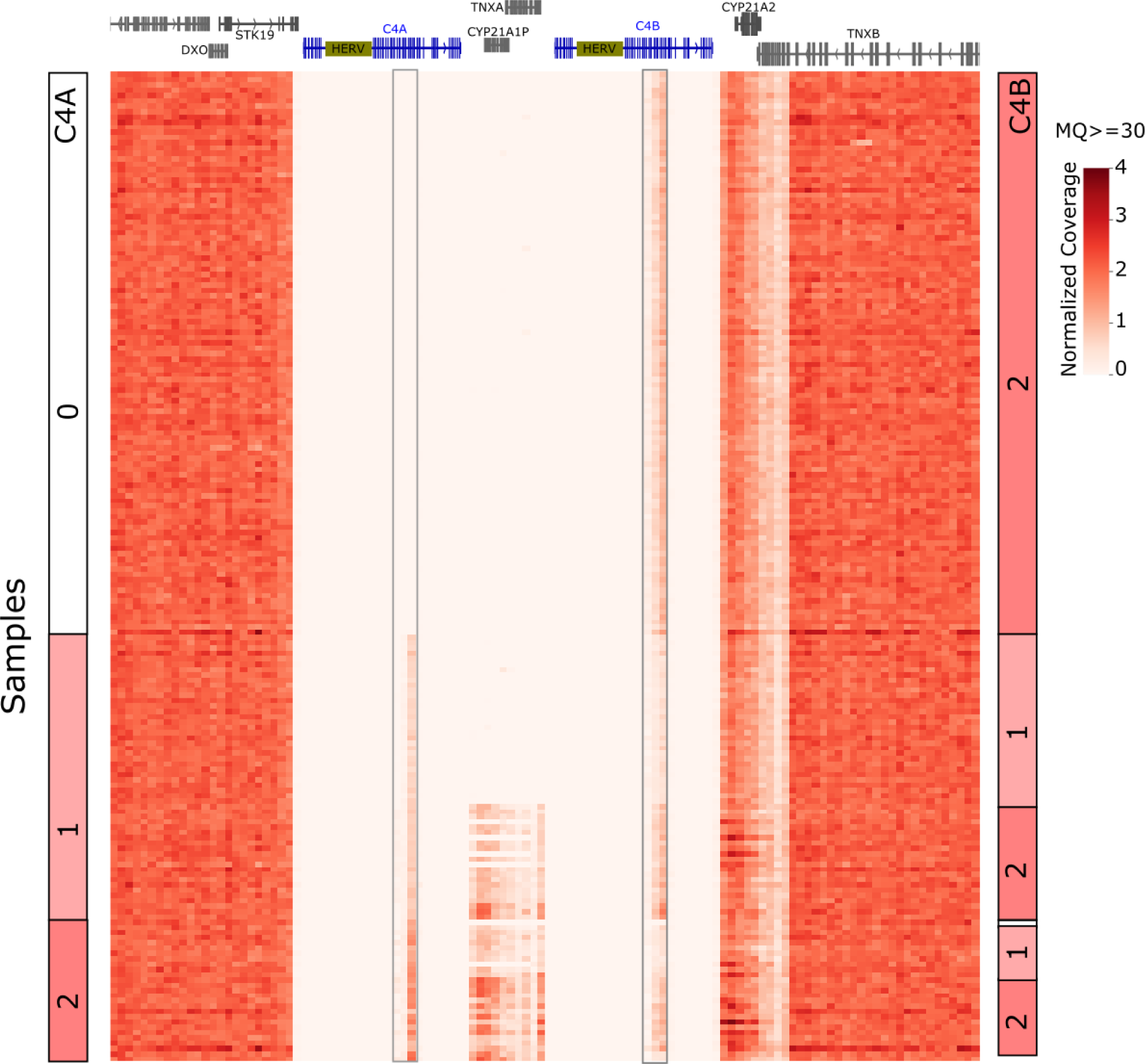
Unique sequence read coverage in C4 region and copy number calls. Uniquely mapping read coverage from WGS data of 188 homozygous DR3-DQ2 individuals. C4A and C4B genes share extensive sequence identity along the genes except a ∼3 kilobase region indicated with boxes. Reads mapping to these regions were used to estimate C4A (left column) and C4B (right column) copy numbers per sample. Samples were sorted based on C4A copy numbers. A maximum value of 4 was used for the heatmap to moderate high outlier values.

## DISCUSSION

We report confirmation of the tri-SNP association with T1D in a much larger, independent cohort. In addition, we show that the 101 tri-SNP haplotype is also associated with reduced risk of developing IA. Although the primary outcome of the TEDDY study was IA, it has been recognized in multiple studies that there may be two distinct “endotypes” of T1D autoimmunity based on the timing of emergence of specific autoantibodies (Battaglia et al., 2020; Johnson et al., 2021; Krischer et al., 2015). One of the proposed endotypes is characterized by the appearance of insulin autoantibody (IAA) early on as the first marker of IA, termed “IAA-first”, and it is associated with HLA DR4-DQ8 haplotype. The second endotype is marked by the emergence at a later time of GAD autoantibodies as the first IA marker, termed “GADA-first”, is associated with DR3-DQ2 haplotype (Krischer et al., 2015). We show that the tri-SNP is significantly associated with protection from the IAA-first outcome but not the GADA-first outcome, indicating that the main T1D protective effect of tri-SNP may be due to delay or prevention of insulin autoantibody generation. This may explain the low incidence of the IAA-first endotype among DR3-DQ2 carrying individuals, given the enrichment of the protective tri-SNP allele on the DR3-DQ2 chromosomes.

Unexpectedly, we discovered that the T1D-protective tri-SNP 101 haplotype is associated with *increased* risk for both autoantibody development (CDA) and clinical CD. This finding is surprising because CD and T1D are thought to share genetic risk factors not only because of the increased co-occurrence of the two pathologies but also because DR3-DQ2 and DR4-DQ8 are major genetic loci for susceptibility for both disorders. An extended HLA-DR3 haplotype (B8) was described in literature as having a differential impact on T1D and CD development in a small sample set of Basque population (Bilbao et al., 2006) but this was not replicated in our sample set. Therefore, the opposite effects of the tri-SNP haplotype on the risk of T1D and CD in our cohort are novel and independent of extended B8 haplotype.

Tri-SNP is a valuable addition to the collection of known genetic and environmental factors that are in use to predict T1D and CD (Bonifacio et al., 2018; Romanos et al., 2014). Knowing whether a high risk HLA child is more likely to develop T1D or CD may aid in decisions about early therapeutic or preventive interventions, such as strict gluten-free diet or preventive anti-CD3 antibody treatment (Herold et al., 2019). Our findings advance our understanding of complex polygenic diseases and should provide a more precise risk/benefit assessment of potential treatments based on data.

Furthermore, we investigated potential mechanisms underlying the differential association of tri-SNP with T1D and CD by analyzing gene expression differences based on the tri-SNP genotype. Complement system genes C4A and C4B emerged as intriguing candidates whose expression changed in opposite direction with respect to tri-SNP genotype. C4 locus has been reported to have frequent copy number variation (Li et al., 2017). Therefore, we explored whether the gene expression changes we observe may be due to changes in gene copy numbers (GCN). Indeed, tri-SNP 101, which is associated with decreased C4A and increased C4B gene expression, was also associated with frequent C4A gene deletions and increased C4B GCN. We also observed that close to 90% of our samples were missing at least one C4 gene. Reduced C4 GCN have been associated with the development of autoimmune diseases such as systemic lupus erythematosus (Pereira et al., 2016), juvenile dermatomyositis (Lintner et al., 2016) and rheumatoid arthritis (Rigby et al., 2012). In addition, lower serum levels of C4 protein have been suggested to predispose to T1D (Vergani et al., 1983). Taken together, reduced total C4 GCN frequent in our high risk HLA cohort is inline with the reports of increased susceptibility of low C4 to autoimmune diseases, and the different disease outcomes based on the tri-SNP haplotype may be, in part, due to the absence of specific C4 genes carried on that haplotype. Although the link between tri-SNP and C4 GCN is intriguing, independent association analyses for C4 genes and diseases were not possible due to the small number of samples with the GCN data. In addition, given the presence of other differentially expressed genes in the MHC locus, it is possible that the differential risk marked by tri-SNP is due to multiple factors.

## Supporting information

TEDDY Study Group Appendix

## Data Availability

All data produced in the present study are available upon reasonable request to the authors

## ACKNOWLEDGMENTS

The TEDDY Study is funded by U01 DK63829, U01 DK63861, U01 DK63821, U01 DK63865, U01 DK63863, U01 DK63836, U01 DK63790, UC4 DK63829, UC4 DK63861, UC4 DK63821, UC4 DK63865, UC4 DK63863, UC4 DK63836, UC4 DK95300, UC4 DK100238, UC4 DK106955, UC4 DK112243, UC4 DK117483, U01 DK124166, U01 DK128847, and Contract No. HHSN267200700014C from the National Institute of Diabetes and Digestive and Kidney Diseases (NIDDK), National Institute of Allergy and Infectious Diseases (NIAID), Eunice Kennedy Shriver National Institute of Child Health and Human Development (NICHD), National Institute of Environmental Health Sciences (NIEHS), Centers for Disease Control and Prevention (CDC), and JDRF. This work is supported in part by the NIH/NCATS Clinical and Translational Science Awards to the University of Florida (UL1 TR000064) and the University of Colorado (UL1 TR002535). The content is solely the responsibility of the authors and does not necessarily represent the official views of the National Institutes of Health.

## Footnotes

1 Based on a receiver-operator curve analysis, prior samples that needed to be re-analyzed with the harmonized assays, included: Denver GADA between −0.015 and 0.042; Bristol GADA between 10.69 and 36.72; Denver IA-2A between −0.004 and 0.016; and Bristol IA-2A between 6.69 and 10.58.

