## Supplementary Information for "Polymorphisms in Intron 1 of HLA-DRA Differentially Associate with Type 1 Diabetes and Celiac Disease and Implicate Involvement of Complement System Genes C4A and C4B"

### SUPPLEMENTARY METHODS

### HLA Inclusion Criteria and Nomenclature Used.

Infants from the general population were eligible for the study if they had any one of the following HLA genotypes (excluding those with DR4 subtype *DRB1*04:03*):

1. *DR3-DQA1*05:01-DQB1*02:01/DR4-DQA1*03:0X-DQB1*03:02*
2. *DR4-DQA1*03:0X-DQB1*03:02/DR4-DQA1*03:0X-DQB1*03:02*
3. *DR4-DQA1*03:0X-DQB1*03:02/DR8-DQA1*04:01-DQB1*04:02*
4. *DR3-DQA1*05:01-DQB1*02:01/DR3-DQA1*05:01-DQB1*02:01*

Infants with a first degree relative with T1D were eligible for enrollment if they had any of the following HLA genotypes:

1. *DR4-DQA1*03:0X-DQB1*03:02/DR3-DQA1*05:01-DQB1*02:01*
2. *DR4-DQA1*03:0X-DQB1*03:02/DR4-DQA1*03:0X-DQB1*03:02*
3. *DR4-DQA1*03:0X-DQB1*03:02/DR8-DQA1*04:01-DQB1*04:02*
4. *DR3-DQA1*05:01-DQB1*02:01/DR3-DQA1*05:01-DQB1*02:01*
5. *DR4-DQA1*03:0X-DQB1*03:02/DR4-DQA1*03:0X-DQB1*02:0X*
6. *DR4-DQA1*03:0X-DQB1*03:02/DR12-DQA1*01:01-DQB1*05:01*
7. *DR4-DQA1*03:0X-DQB1*03:02/DR13-DQA1*01:02-DQB1*06:04*
8. *DR4-DQA1*03:0X-DQB1*03:02/DR4-DQA1*03:0X-DQB1*03:04*
9. *DR4-DQA1*03:0X-DQB1*03:02/DR9-DQA1*03:0X-DQB1*03:03*
10. *DR3-DQA1*05:01-DQB1*02:01/DR9-DQA1*03:0X-DQB1*03:03*

Explanation of HLA nomenclature used was as follows.

DR3 = *DRB1*03:01*; DR4 = *DRB1*04:01, *04:02, *04:04, *04:05,* or **04:07*; *DQA1*03:0X* = any *DQA1*03*; *DQB1*03:02* = *DQB1*03:02* or *DQB1*03:04*; *DQB1*02:0X* = any *DQB1*02*; DR8 = any *DRB1*08*; DR9 = *DRB1*09:01*; DR12 = *any DRB1*12*; DR13 = any *DRB1*13.*

### Total C4 Gene Copy Number (GCN) estimation

We estimated the total C4 GCN based on the read coverage of the C4 region including C4A and C4B but excluding the intronic HERV insertion (Supplementary Figure 13A). Histograms of per sample coverage for C4 region as well as 25 kilobases flanking regions on both sides were plotted (Supplementary Figure 13 B). While the flanking regions showed a single peak around 2, indicating a normal diploid status; C4 region showed 3 peaks around 1, 1.5 and 2, likely representing 2, 3 and 4 GCNs, respectively. Since the three peaks in the C4 histogram was well separated, we estimated total C4 GCN based on these peaks: samples under the first peak (coverage < 1.4) were assigned 2 GCN; those under the second peak (1.4<coverage<1.8) were assigned 3 GCN and those under the third peak (coverage > 1.8) were assigned 4 GCN.

### C4A and C4B Gene Copy Number (GCN) estimation

Histograms of coverage based on reads uniquely mapping to either C4A or C4B were created (Supplementary Figure 13 C). We observed a major peak at 0 coverage for C4A, indicating total C4A deletions and a separate peak around 0.3 indicating 1 GCN; and 2 peaks for C4B around 0.3 and 0.6, which likely indicate GCNs of 1 and 2. Since total gene deletions were well separated in the histograms for C4A and C4B, we first assigned the GCNs of C4A or C4B null samples (average coverage below 0.04). However, because the peaks for 1 and 2 GCNs were not clearly separated, the rest of the C4A and C4B GCNs were estimated based on the total C4 GCN and the ratio of unique coverage of C4A and C4B. The exact heuristic used was: if a sample was C4A null, C4B was assigned the same value as total C4 GCN; if C4B null, C4A was assigned the C4 GCN. For samples where neither C4A nor C4B was null: if C4 GCN was 2, C4A and C4B each were assigned 1 GCN; if C4 GCN was 3, C4A and C4B coverage were compared and larger received 2 GCN and smaller 1 GCN; if C4 GCN was 4, C4A and C4B were each assigned 2 GCN unless the coverage ratio between them was larger than 2.5 in which case the gene with more coverage was assigned 3 and the other 1 GCN.

### SUPPLEMENTARY TEXT

### Impact of known risk factors on T1D diagnosis

Known risk factors HLA DR3/DR4 genotype (HR=3.20, CI=2.18-4.70, p=3.16e-9) and having an affected FDR (HR=3.11, CI=2.42-3.98, p=3.75e-19) were the largest risk factors, as expected (Krischer et al., 2017; Sharma et al., 2018) (Figure 1A, Supplementary Figure 7, Supplementary Data). All six previously identified GWAS associations with T1D were also significantly associated with T1D diagnosis (Krischer et al., 2017; Sharma et al., 2018) . While genetic ancestry and sex were not detected as significant risk factors, subjects from Finland were marginally at higher risk (HR=1.34, p=0.04, Supplementary Figure 1, Supplementary Data).

Among DR3 homozygous individuals, FDR (HR=2.86, CI=1.22-6.67, p=0.02) and rs2476601 (PTPN22, HR=2.51, CI=1.38-4.56, p=0.003) remained significant risk factors (Supplementary Figure 2).

### Impact of known risk factors on islet autoantibody (IA) development

Among the controlled covariates, having a first degree relative (FDR) with T1D (HR=2.16, CI=1.78-2.62, p=5.76e-15, Figure 1 B), HLA DR3/DR4 genotype (HR=1.83, CI=1.41-2.39, p=6.96e-6, Supplementary Figure 3) were significant risk factors for IA; and male subjects were found to be at marginally higher risk of developing IA (HR=1.19, CI=1.03-1.36, p=0.01), consistent with previous TEDDY reports (Krischer et al., 2017; Sharma et al., 2018).

Subjects from Finland (HR=1.34, CI=1.11-1.63, p=0.002) and Sweden (HR=1.25, CI=1.06-1.48, p=0.008) were at slightly higher risk than subjects from the United States (Supplementary Figure 3). Seven GWAS loci reported to be associated with IA in the TEDDY cohort (Krischer et al., 2017; Sharma et al., 2018) were confirmed to be significantly associated and in the same direction (protective vs risk) with IA development in our analysis.

When analysis was restricted to DR3 homozygous individuals, having an FDR with T1D (HR=2.42, CI=1.49-3.93, p=3.37e-4, Figure 1 B) remained a significant risk factor within this group, but other controlled covariates were no longer significant (Supplementary Figure 4).

### SUPPLEMENTARY TABLES

**Supplementary Table 1.** Descriptive characteristics of children with respect to islet autoimmunity (IA) outcome in number (percentage).

|  | **IA** | | |
| --- | --- | --- | --- |
|  | **No** | **Yes** | **All** |
| **Sex** |  |  |  |
| **Female** | 3388 (49.5) | 390 (45.5) | 3778 (49.0) |
| **Male** | 3457 (50.5) | 468 (54.5) | 3925 (51.0) |
| **POP** |  |  |  |
| **EUR** | 6122 (89.4) | 796 (92.8) | 6918 (89.8) |
| **AMR** | 647 (9.5) | 56 (6.5) | 703 (9.1) |
| **AFR** | 76 (1.1) | 6 (0.7) | 82 (1.1) |
| **HLA Type** |  |  |  |
| **DR1/DR4** | 142 (2.1) | 18 (2.1) | 160 (2.1) |
| **DR3/DR3** | 1486 (21.7) | 121 (14.1) | 1607 (20.9) |
| **DR3/DR4** | 2599 (38.0) | 418 (48.7) | 3017 (39.2) |
| **DR4/DR13** | 47 (0.7) | 10 (1.2) | 57 (0.7) |
| **DR4/DR4** | 1371 (20.0) | 158 (18.4) | 1529 (19.8) |
| **DR4/DR8** | 1200 (17.5) | 133 (15.5) | 1333 (17.3) |
| **Country** |  |  |  |
| **US** | 2885 (42.1) | 297 (34.6) | 3182 (41.3) |
| **SWE** | 2019 (29.5) | 290 (33.8) | 2309 (30.0) |
| **FIN** | 1482 (21.7) | 214 (24.9) | 1696 (22.0) |
| **GER** | 459 (6.7) | 57 (6.6) | 516 (6.7) |
| **FDR** |  |  |  |
| **0** | 6155 (89.9) | 691 (80.5) | 6846 (88.9) |
| **1** | 690 (10.1) | 167 (19.5) | 857 (11.1) |
| **All** | 6845 (88.9) | 858 (11.1) | 7703 (100) |

**Supplementary Table 2.** Descriptive characteristics of children with respect to type 1 diabetes diagnosis (T1D) outcome in number (percentage).

|  | **T1D** | | |
| --- | --- | --- | --- |
| **Sex** | **No** | **Yes** | **All** |
| Female | 3597 (49.2) | 181 (45.6) | 3778 (49.0) |
| Male | 3709 (50.8) | 216 (54.4) | 3925 (51.0) |
| **POP** |  |  |  |
| EUR | 6552 (89.7) | 366 (92.2) | 6918 (89.8) |
| AMR | 674 (9.2) | 29 (7.3) | 703 (9.1) |
| AFR | 80 (1.1) | 2 (0.5) | 82 (1.1) |
| **HLA Type** |  |  |  |
| DR1/DR4 | 146 (2.0) | 14 (3.5) | 160 (2.1) |
| DR3/DR3 | 1571 (21.5) | 36 (9.1) | 1607 (20.9) |
| DR3/DR4 | 2797 (38.3) | 220 (55.4) | 3017 (39.2) |
| DR4/DR13 | 51 (0.7) | 6 (1.5) | 57 (0.7) |
| DR4/DR4 | 1457 (19.9) | 72 (18.1) | 1529 (19.8) |
| DR4/DR8 | 1284 (17.6) | 49 (12.3) | 1333 (17.3) |
| **Country** |  |  |  |
| US | 3034 (41.5) | 148 (37.3) | 3182 (41.3) |
| SWE | 2204 (30.2) | 105 (26.4) | 2309 (30.0) |
| FIN | 1591 (21.8) | 105 (26.4) | 1696 (22.0) |
| GER | 477 (6.5) | 39 (9.8) | 516 (6.7) |
| **FDR** |  |  |  |
| 0 | 6560 (89.8) | 286 (72.0) | 6846 (88.9) |
| 1 | 746 (10.2) | 111 (28.0) | 857 (11.1) |
| **All** | 7306 (94.8) | 397 (5.2) | 7703 (100) |

**Supplementary Table 3.** Descriptive characteristics of children with respect to GADA-first appearing antibody outcome in number (percentage).

|  | **GADA first** | | |
| --- | --- | --- | --- |
|  | **No** | **Yes** | **All** |
| **Sex** |  |  |  |
| Female | 3601 (49.2) | 177 (46.3) | 3778 (49.0) |
| Male | 3720 (50.8) | 205 (53.7) | 3925 (51.0) |
| **POP** |  |  |  |
| EUR | 6565 (89.7) | 353 (92.4) | 6918 (89.8) |
| AMR | 678 (9.3) | 25 (6.5) | 703 (9.1) |
| AFR | 78 (1.1) | 4 (1.0) | 82 (1.1) |
| **HLA Type** |  |  |  |
| DR1/DR4 | 157 (2.1) | 3 (0.8) | 160 (2.1) |
| DR3/DR3 | 1522 (20.8) | 85 (22.3) | 1607 (20.9) |
| DR3/DR4 | 2828 (38.6) | 189 (49.5) | 3017 (39.2) |
| DR4/DR13 | 55 (0.8) | 2 (0.5) | 57 (0.7) |
| DR4/DR4 | 1472 (20.1) | 57 (14.9) | 1529 (19.8) |
| DR4/DR8 | 1287 (17.6) | 46 (12.0) | 1333 (17.3) |
| **Country** |  |  |  |
| US | 3036 (41.5) | 146 (38.2) | 3182 (41.3) |
| SWE | 2164 (29.6) | 145 (38.0) | 2309 (30.0) |
| FIN | 1622 (22.2) | 74 (19.4) | 1696 (22.0) |
| GER | 499 (6.8) | 17 (4.5) | 516 (6.7) |
| **FDR** |  |  |  |
| No | 6531 (89.2) | 315 (82.5) | 6846 (88.9) |
| Yes | 790 (10.8) | 67 (17.5) | 857 (11.1) |
| **All** | 7321 (95.0) | 382 (5.0) | 7703 (100) |

**Supplementary Table 4.** Descriptive characteristics of children with respect to IAA-first appearing antibody outcome in number (percentage).

|  | **IAA first** | | |
| --- | --- | --- | --- |
|  | **No** | **Yes** | **All** |
| **Sex** |  |  |  |
| Female | 3636 (49.2) | 142 (45.4) | 3778 (49.0) |
| Male | 3754 (50.8) | 171 (54.6) | 3925 (51.0) |
| **POP** |  |  |  |
| EUR | 6624 (89.6) | 294 (93.9) | 6918 (89.8) |
| AMR | 685 (9.3) | 18 (5.8) | 703 (9.1) |
| AFR | 81 (1.1) | 1 (0.3) | 82 (1.1) |
| **HLA Type** |  |  |  |
| DR1/DR4 | 151 (2.0) | 9 (2.9) | 160 (2.1) |
| DR3/DR3 | 1578 (21.4) | 29 (9.3) | 1607 (20.9) |
| DR3/DR4 | 2870 (38.8) | 147 (47.0) | 3017 (39.2) |
| DR4/DR13 | 52 (0.7) | 5 (1.6) | 57 (0.7) |
| DR4/DR4 | 1471 (19.9) | 58 (18.5) | 1529 (19.8) |
| DR4/DR8 | 1268 (17.2) | 65 (20.8) | 1333 (17.3) |
| **Country** |  |  |  |
| US | 3082 (41.7) | 100 (31.9) | 3182 (41.3) |
| SWE | 2215 (30.0) | 94 (30.0) | 2309 (30.0) |
| FIN | 1597 (21.6) | 99 (31.6) | 1696 (22.0) |
| GER | 496 (6.7) | 20 (6.4) | 516 (6.7) |
| **FDR** |  |  |  |
| No | 6600 (89.3) | 246 (78.6) | 6846 (88.9) |
| Yes | 790 (10.7) | 67 (21.4) | 857 (11.1) |
| **All** | 7390 (95.9) | 313 (4.1) | 7703 (100) |

**Supplementary Table 5.** Descriptive characteristics of children with respect to celiac disease diagnosis (CD) outcome in number (percentage).

|  | **CD** | | |
| --- | --- | --- | --- |
|  | **No** | **Yes** | **All** |
| **Sex** |  |  |  |
| Female | 3413 (48.2) | 365 (59.2) | 3778 (49.0) |
| Male | 3673 (51.8) | 252 (40.8) | 3925 (51.0) |
| **POP** |  |  |  |
| EUR | 6325 (89.3) | 593 (96.1) | 6918 (89.8) |
| AMR | 681 (9.6) | 22 (3.6) | 703 (9.1) |
| AFR | 80 (1.1) | 2 (0.3) | 82 (1.1) |
| **HLA Type** |  |  |  |
| DR1/DR4 | 155 (2.2) | 5 (0.8) | 160 (2.1) |
| DR3/DR3 | 1308 (18.5) | 299 (48.5) | 1607 (20.9) |
| DR3/DR4 | 2805 (39.6) | 212 (34.4) | 3017 (39.2) |
| DR4/DR13 | 54 (0.8) | 3 (0.5) | 57 (0.7) |
| DR4/DR4 | 1444 (20.4) | 85 (13.8) | 1529 (19.8) |
| DR4/DR8 | 1320 (18.6) | 13 (2.1) | 1333 (17.3) |
| **Country** |  |  |  |
| US | 2955 (41.7) | 227 (36.8) | 3182 (41.3) |
| SWE | 2049 (28.9) | 260 (42.1) | 2309 (30.0) |
| FIN | 1594 (22.5) | 102 (16.5) | 1696 (22.0) |
| GER | 488 (6.9) | 28 (4.5) | 516 (6.7) |
| **FDR** |  |  |  |
| No | 6296 (88.9) | 550 (89.1) | 6846 (88.9) |
| Yes | 790 (11.1) | 67 (10.9) | 857 (11.1) |
| **All** | 7086 (92.0) | 617 (8.0) | 7703 (100) |

**Supplementary Table 6.** Descriptive characteristics of children with respect to celiac disease autoimmunity (CDA) outcome in number (percentage).

|  | **CDA** | | |
| --- | --- | --- | --- |
|  | **No** | **Yes** | **All** |
| **Sex** |  |  |  |
| Female | 2555 (47.2) | 735 (56.8) | 3290 (49.0) |
| Male | 2860 (52.8) | 559 (43.2) | 3419 (51.0) |
| **POP** |  |  |  |
| EUR | 4855 (89.7) | 1237 (95.6) | 6092 (90.8) |
| AMR | 515 (9.5) | 55 (4.3) | 570 (8.5) |
| AFR | 45 (0.8) | 2 (0.2) | 47 (0.7) |
| **HLA Type** |  |  |  |
| DR1/DR4 | 134 (2.5) | 8 (0.6) | 142 (2.1) |
| DR3/DR3 | 870 (16.1) | 530 (41.0) | 1400 (20.9) |
| DR3/DR4 | 2146 (39.6) | 503 (38.9) | 2649 (39.5) |
| DR4/DR13 | 46 (0.8) | 4 (0.3) | 50 (0.7) |
| DR4/DR4 | 1130 (20.9) | 193 (14.9) | 1323 (19.7) |
| DR4/DR8 | 1089 (20.1) | 56 (4.3) | 1145 (17.1) |
| **Country** |  |  |  |
| US | 2232 (41.2) | 471 (36.4) | 2703 (40.3) |
| SWE | 1588 (29.3) | 479 (37.0) | 2067 (30.8) |
| FIN | 1262 (23.3) | 272 (21.0) | 1534 (22.9) |
| GER | 333 (6.1) | 72 (5.6) | 405 (6.0) |
| **FDR** |  |  |  |
| No | 4794 (88.5) | 1147 (88.6) | 5941 (88.6) |
| Yes | 621 (11.5) | 147 (11.4) | 768 (11.4) |
| **All** | 5415 (80.7) | 1294 (19.3) | 6709 (100) |

**Supplementary Table 7. Previously published GWAS associations.** SNPs used as covariates in our CoxPH analysis. CD, CDA, T1D and IA columns indicating whether the SNP has been shown to be associated with that outcome and hence used in the model (yes) or not (no). Statistically significant hazard ratios for the associated outcome are also provided under HR columns, protective associations in bold.

| **SNP** | **Locus** | **CD** | **CDA** | **T1D** | **IA** | **Publication** | **HR CD** | **HR CDA** | **HR T1D** | **HR IA** |
| --- | --- | --- | --- | --- | --- | --- | --- | --- | --- | --- |
| rs4851575 | IL18R1, IL18RAP | yes | no | no | no | (Sharma et al., 2016) | 1.45 |  |  |  |
| rs114569351 | PLEK,  FBXO48 | yes | no | no | no | (Sharma et al., 2016) | 2.64 |  |  |  |
| rs12493471 | CCR9,  LZTFL1,  CXCR6 | yes | no | no | no | (Sharma et al., 2016) | 1.40 |  |  |  |
| rs1054091 | RSPH3,  TAGAP | yes | no | no | no | (Sharma et al., 2016) | 1.59 |  |  |  |
| rs72704176 | ASH1L | yes | no | no | no | (Sharma et al., 2016) | 2.26 |  |  |  |
| rs3771689 | BAZ2B | yes | no | no | no | (Sharma et al., 2016) | **0.56** |  |  |  |
| rs13014907 | ZNF804A | yes | no | no | no | (Sharma et al., 2016) | 2.46 |  |  |  |
| rs11739460 | TCOF1 | yes | no | no | no | (Sharma et al., 2016) | 1.41 |  |  |  |
| rs77532435 | GRB10 | yes | no | no | no | (Sharma et al., 2016) | 2.05 |  |  |  |
| rs6967298 | AUTS2 | yes | no | no | no | (Sharma et al., 2016) | **0.61** |  |  |  |
| rs61751041 | LAMB1 | yes | no | no | no | (Sharma et al., 2016) | 2.23 |  |  |  |
| rs2409747 | XKR6 | yes | yes | no | no | (Sharma et al., 2016) | 1.58 | 1.37 |  |  |
| rs12990970 | NPM1P33,  CTLA4 | no | yes | no | no | (Sharma et al., 2016) |  | **0.76** |  |  |
| rs11709472 | LPP | no | yes | no | no | (Sharma et al., 2016) |  | **0.80** |  |  |
| rs72717025 | FCGR2A | no | yes | no | no | (Sharma et al., 2016) |  | 1.84 |  |  |
| rs114157400 | BANK1 | no | yes | no | no | (Sharma et al., 2016) |  | 1.62 |  |  |
| rs117561283 | IFNG | no | yes | no | no | (Sharma et al., 2016) |  | 1.81 |  |  |
| rs8013918 | FOS | no | yes | no | no | (Sharma et al., 2016) |  | **0.80** |  |  |
| rs73043122 | RNASET2,  MIR3939 | no | no | yes | no | (Sharma et al., 2018) |  |  | 3.35 |  |
| rs113306148 | PLEKHA1,  MIR3941 | no | no | yes | no | (Sharma et al., 2018) |  |  | 3.06 |  |
| rs428595 | PPIL2 | no | no | yes | yes | (Sharma et al., 2018) |  |  | 3.42 | 2.46 |
| rs1004446 | INS | no | no | yes | yes | (Krischer et al., 2017) |  |  | **0.55** | **0.67** |
| rs2476601 | PTPN22 | no | no | yes | yes | (Krischer et al., 2017) |  |  | 1.91 | 1.73 |
| rs2292239 | ERBB3 | no | no | yes | yes | (Krischer et al., 2017) |  |  | 1.68 | 1.45 |
| rs3184504 | SH2B3 | no | no | no | yes | (Krischer et al., 2017) |  |  |  | 1.40 |
| rs9934817 | RBFOX1 | no | no | no | yes | (Sharma et al., 2018) |  |  |  | 2.66 |
| rs11705721 | PXK, PDHB | no | no | no | yes | (Sharma et al., 2018) |  |  |  | 1.41 |

**Supplementary Table 8.** Estimated C4A gene copy number with respect to tri-SNP haplotype.

|  | **tri-SNP 101** | | |
| --- | --- | --- | --- |
| **C4A Copy Number** | **0** | **1** | **2** |
| **0** | 0 | 0 | 107 |
| **1** | 0 | 44 | 10 |
| **2** | 9 | 9 | 7 |
| **3** | 1 | 0 | 1 |

**Supplementary Table 9.** Estimated C4B gene copy number with respect to tri-SNP haplotype.

|  | **tri-SNP 101** | | |
| --- | --- | --- | --- |
| **C4B Copy Number** | **0** | **1** | **2** |
| **0** | 1 | 0 | 0 |
| **1** | 5 | 34 | 4 |
| **2** | 4 | 17 | 119 |
| **3** | 0 | 2 | 2 |

### SUPPLEMENTARY FIGURES

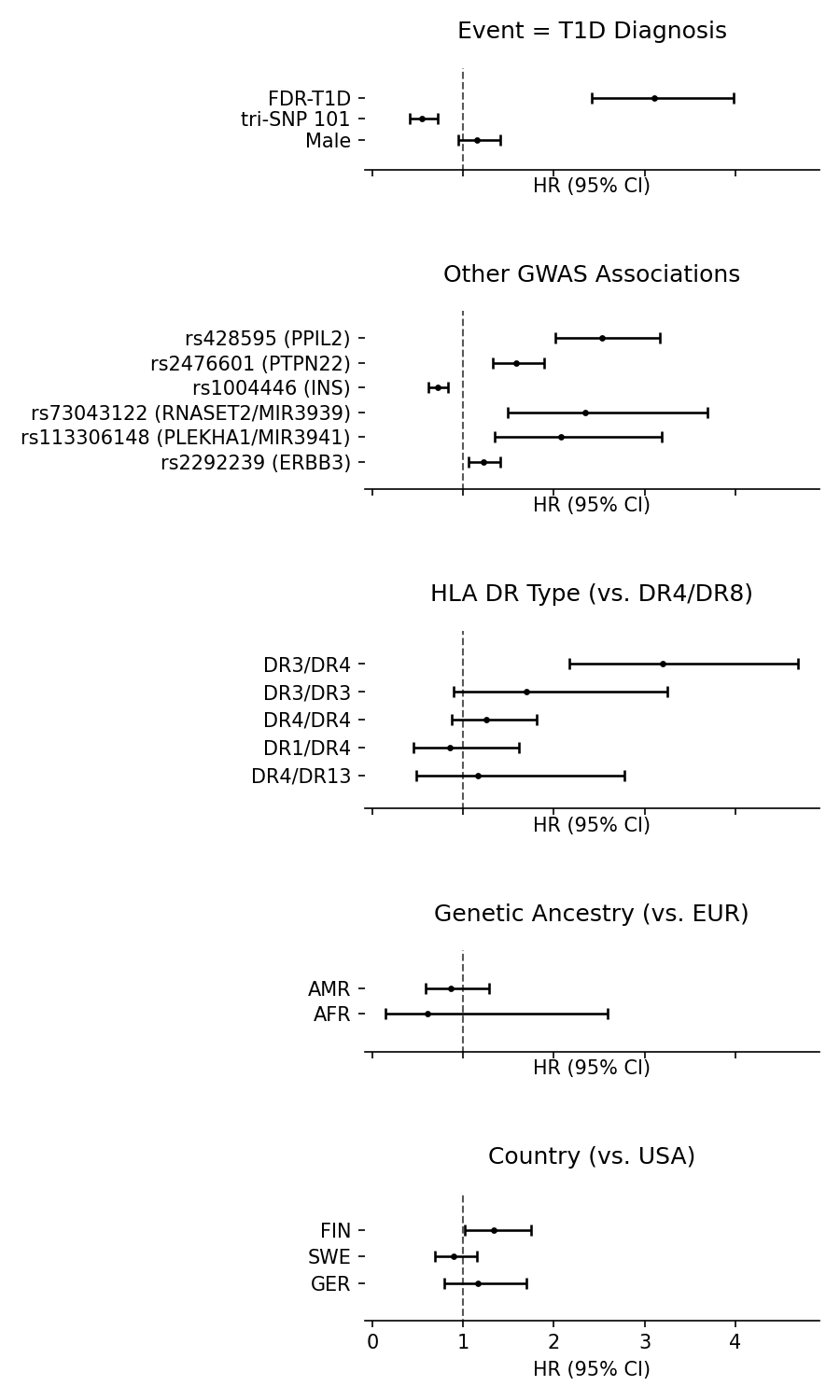

**Supplementary Figure 1.** Complete model output for T1D outcome. HR values and 95% CI are plotted for each covariate included in the model. Vertical line marks the HR=1 (no change in risk). N_observations_= 7638, N_events_= 395.

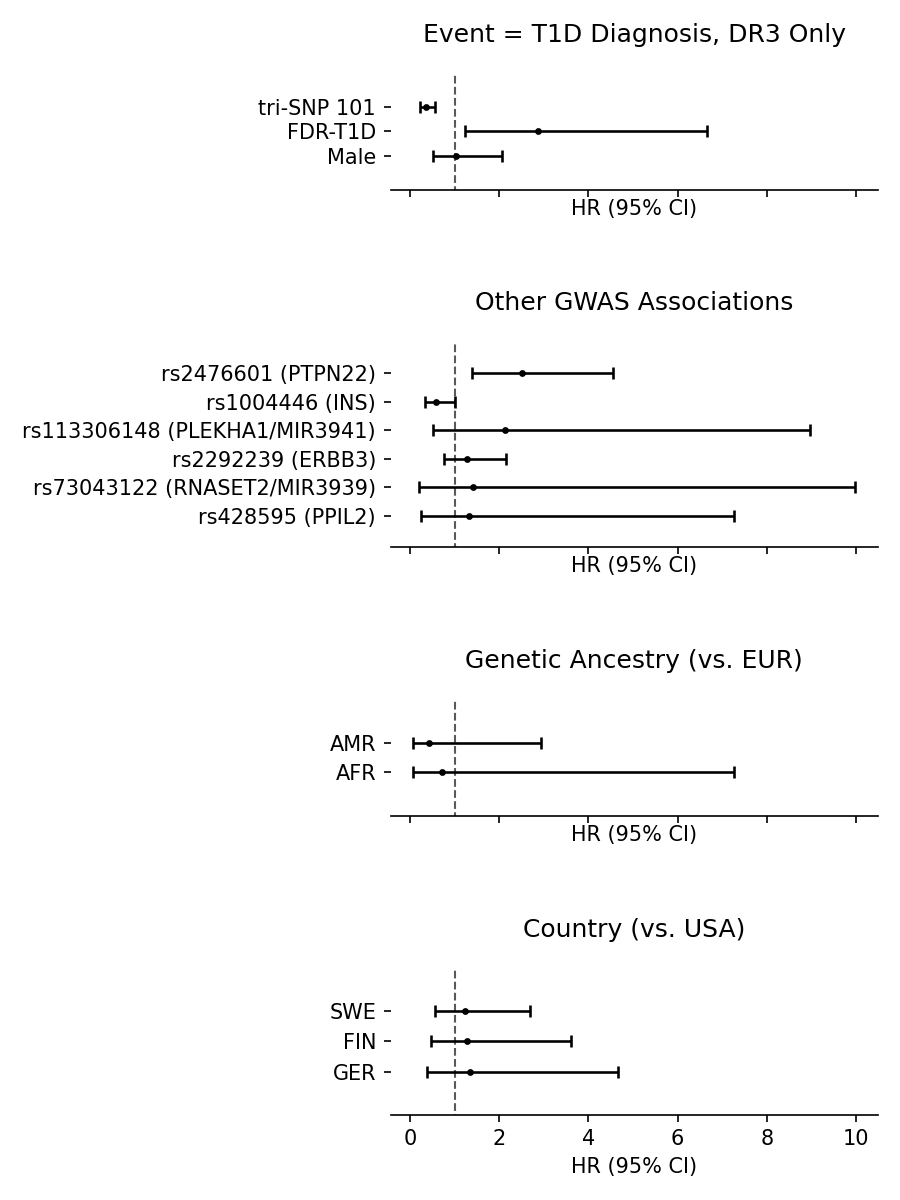

**Supplementary Figure 2.** Complete model output for T1D outcome using only DR3-DQ2 homozygotes. HR values and 95% CI are plotted for each covariate included in the model. Vertical line marks the HR=1 (no change in risk). N_observations_= 1589, N_events_= 36.

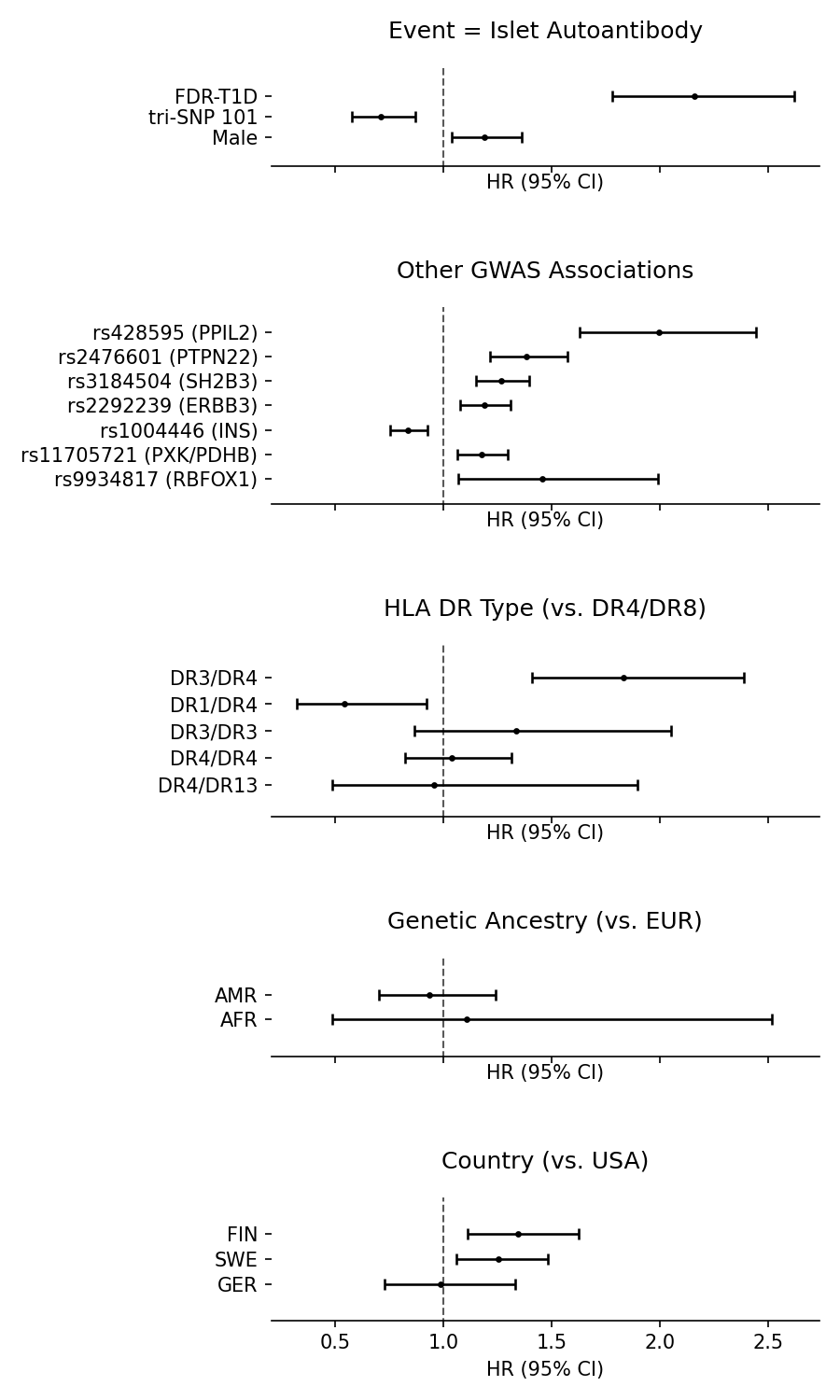

**Supplementary Figure 3.** Complete model output for IA outcome using all samples. HR values and 95% CI are plotted for each covariate included in the model. Vertical line marks the HR=1 (no change in risk). N_observations_= 7614, N_events_= 855.

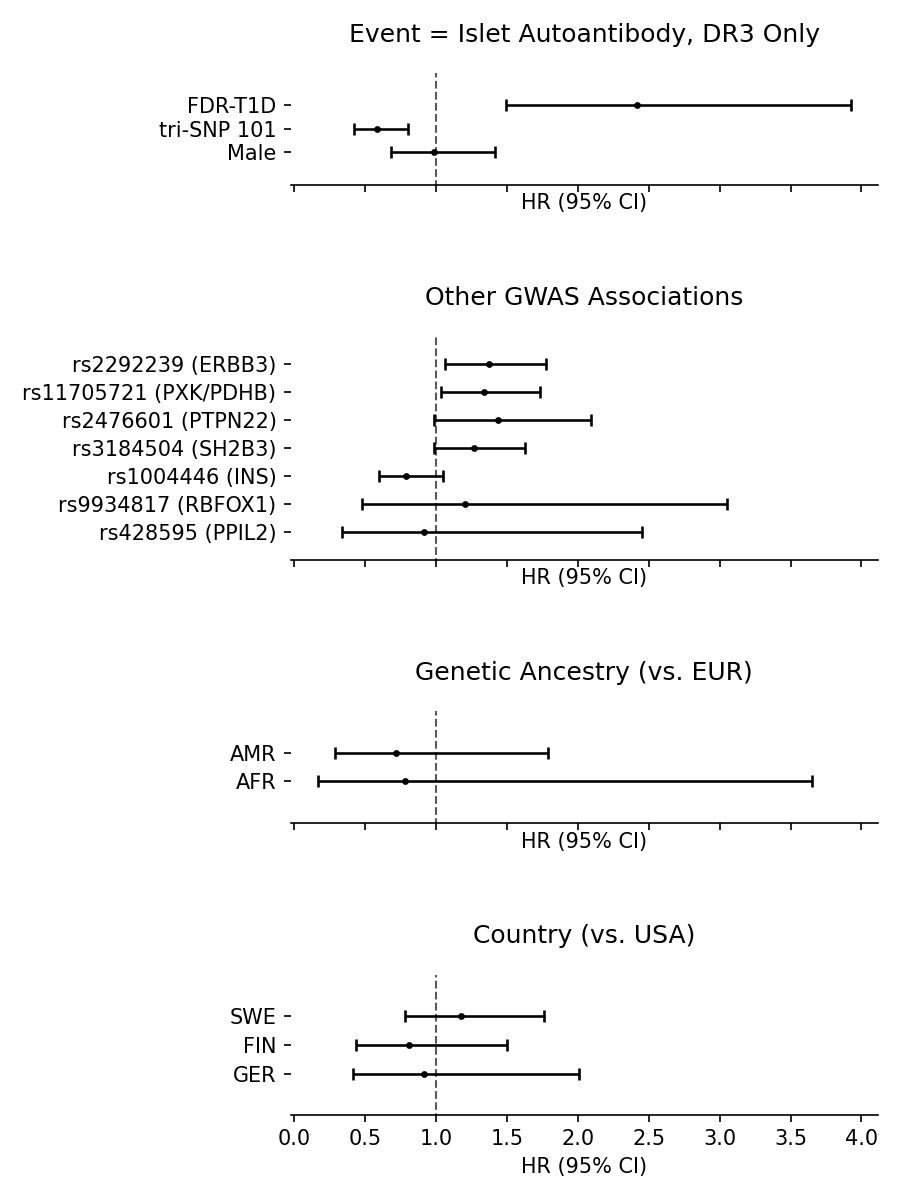

**Supplementary Figure 4.** Complete model output for IA outcome using only DR3-DQ2 homozygotes. HR values and 95% CI are plotted for each covariate included in the model. Vertical line marks the HR=1 (no change in risk). N_observations_= 1585, N_events_= 121.

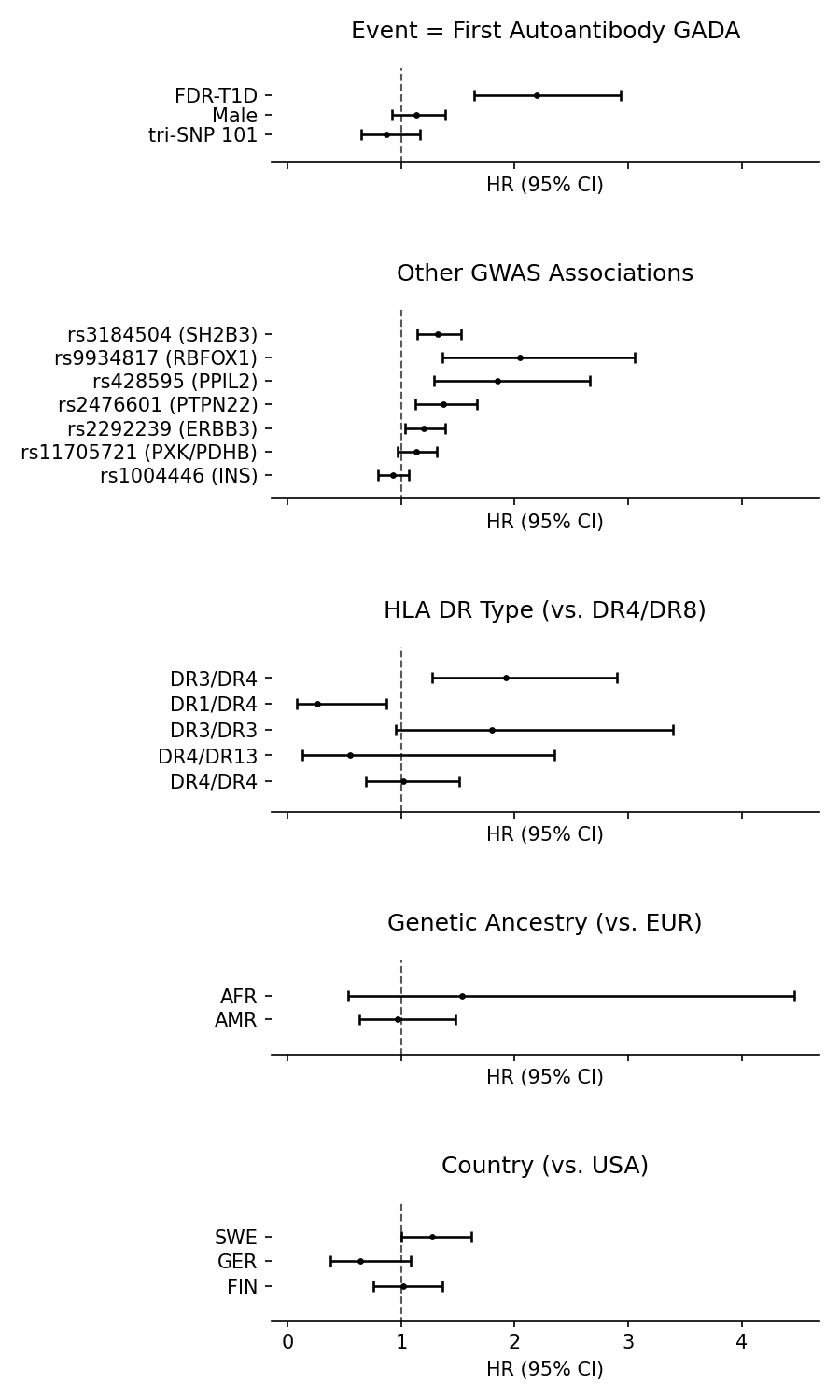

**Supplementary Figure 5.** Complete model output for GADA-first outcome. HR values and 95% CI are plotted for each covariate included in the model. Vertical line marks the HR=1 (no change in risk). N_observations_= 7614, N_events_= 382.

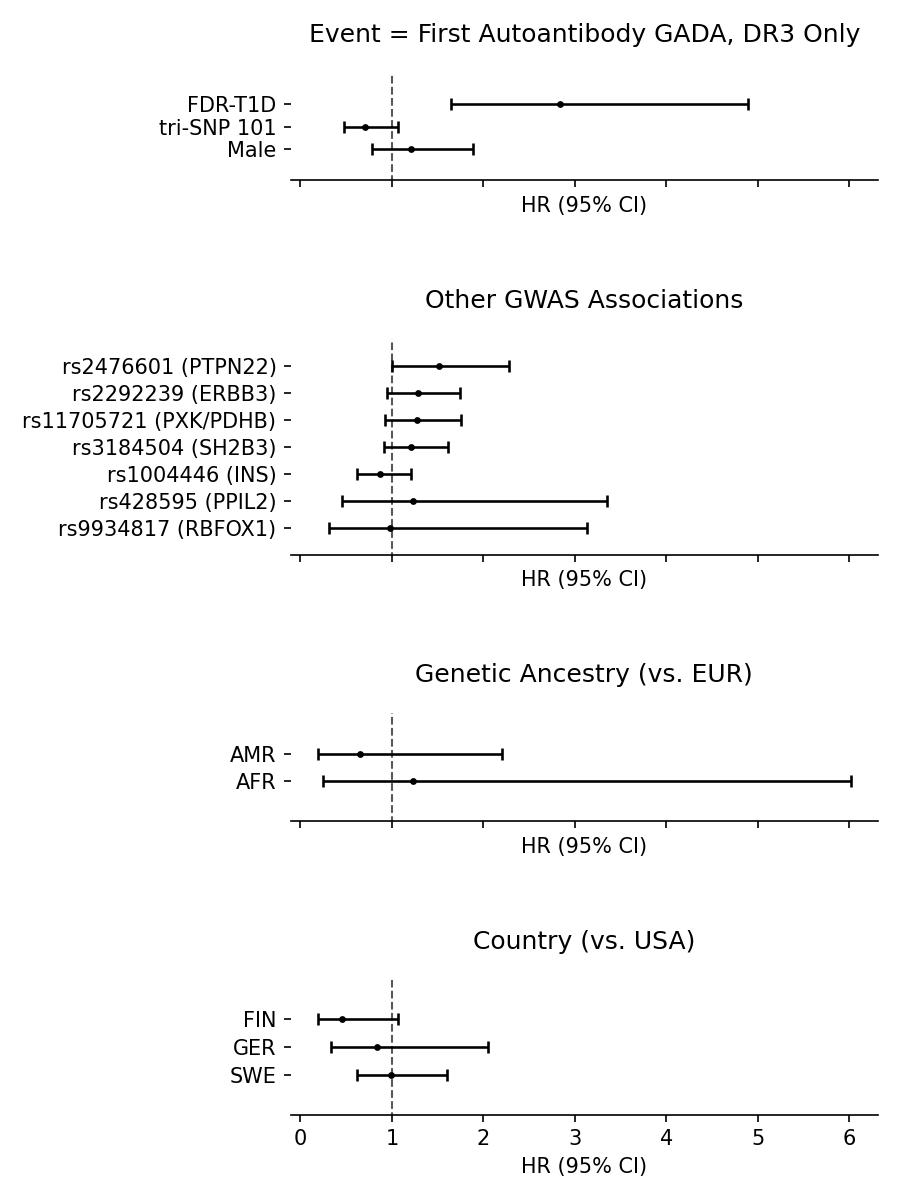

**Supplementary Figure 6.** Complete model output for GADA-first outcome using only DR3-DQ2 homozygotes. HR values and 95% CI are plotted for each covariate included in the model. Vertical line marks the HR=1 (no change in risk). N_observations_= 1585, N_events_= 85.

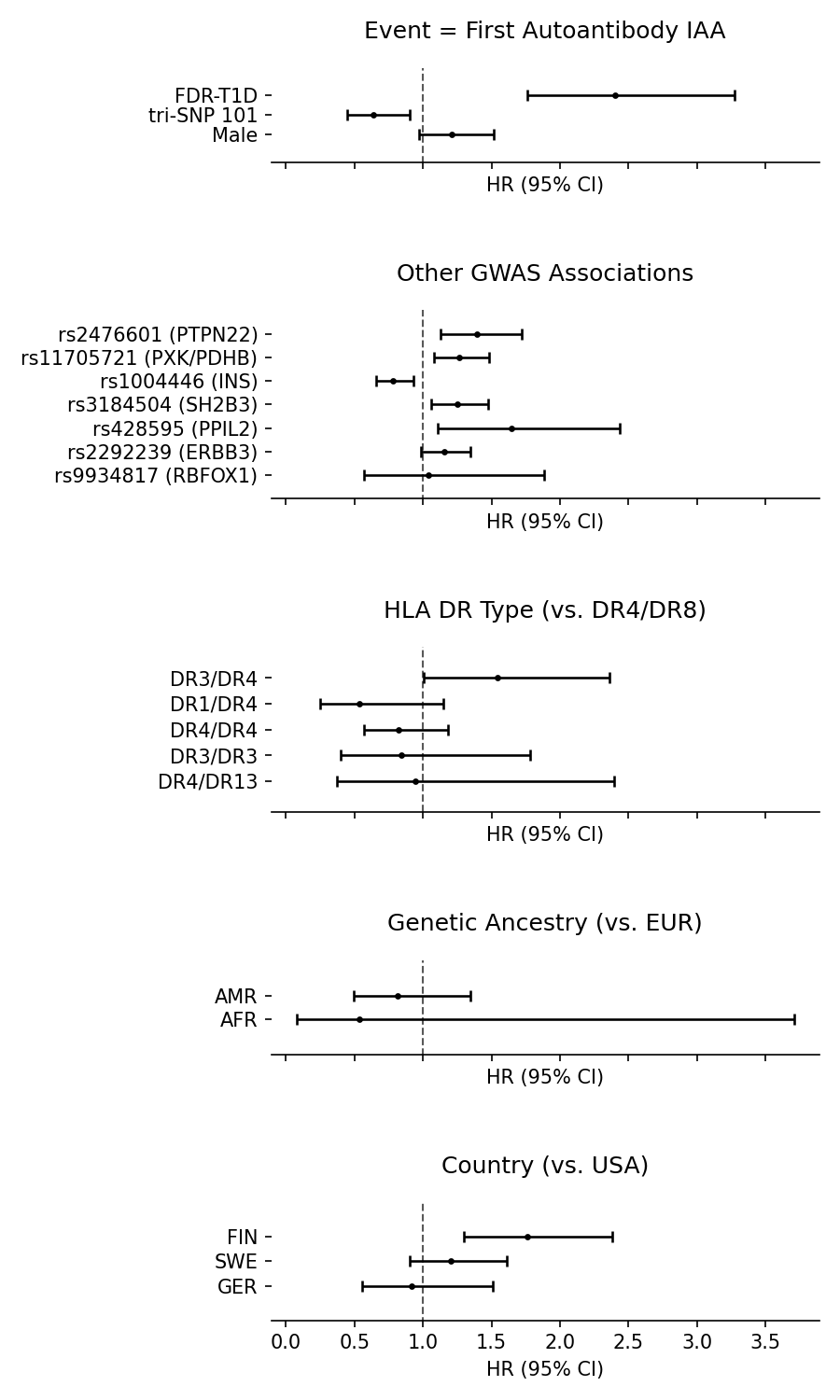

**Supplementary Figure 7.** Complete model output for IAA-first outcome. HR values and 95% CI are plotted for each covariate included in the model. Vertical line marks the HR=1 (no change in risk). N_observations_= 7614, N_events_= 313.

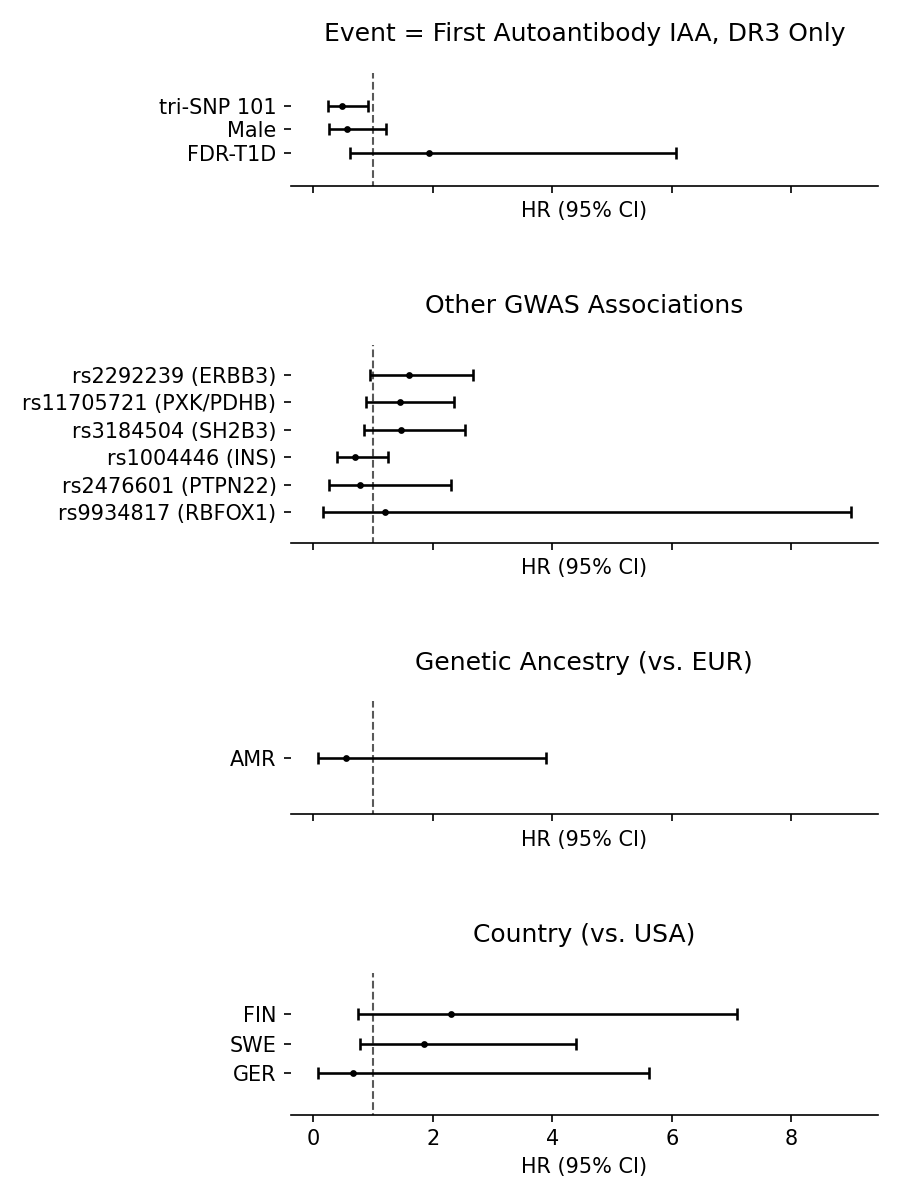

**Supplementary Figure 8.** Complete model output for IAA-first outcome using only DR3-DQ2 homozygotes. HR values and 95% CI are plotted for each covariate included in the model. Vertical line marks the HR=1 (no change in risk). N_observations_= 1570, N_events_= 29.

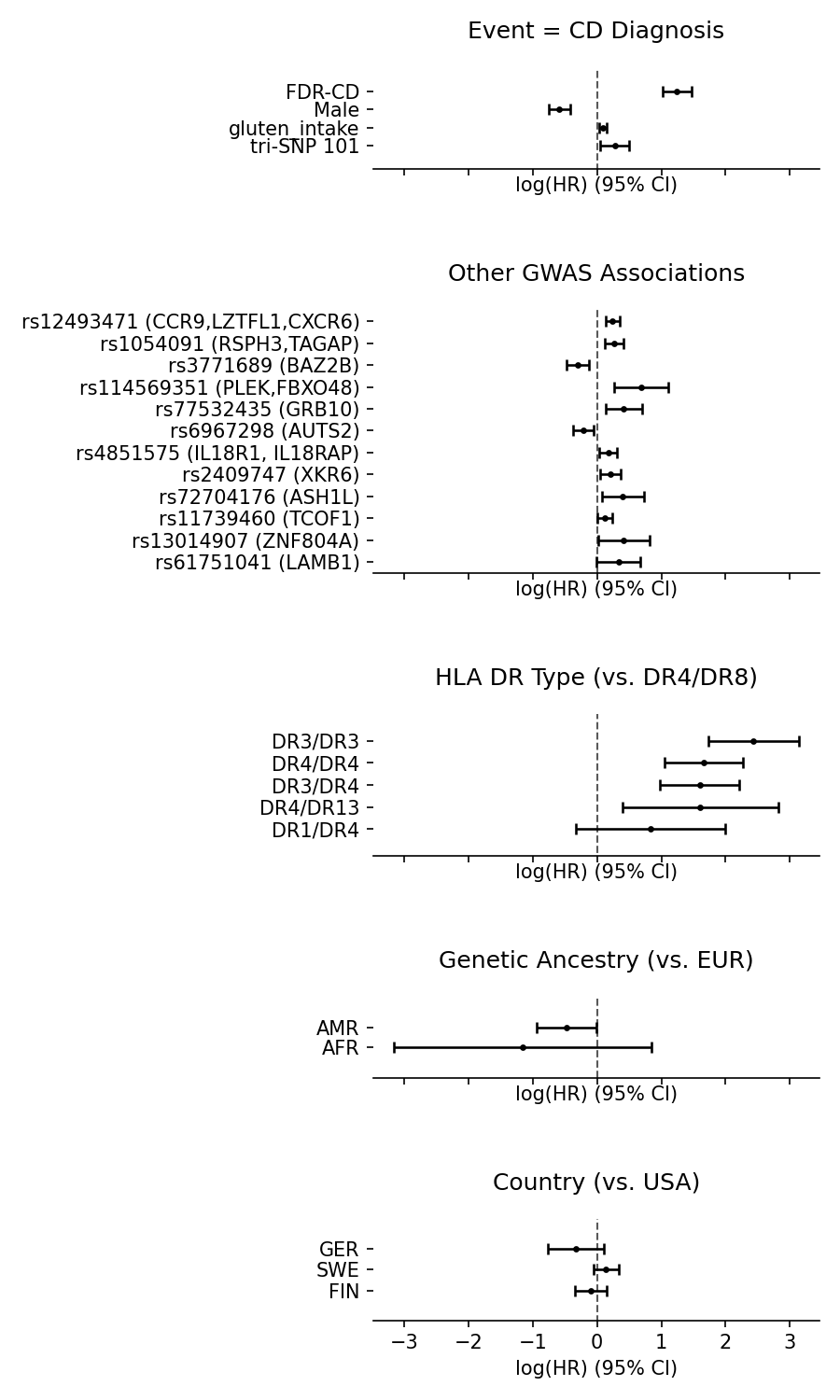

**Supplementary Figure 9.** Complete model output for CD outcome. log(HR) values and 95% CI are plotted for each covariate included in the model. Vertical line marks the log(HR)=0 (no change in risk). N_observations_= 6530, N_events_= 608.

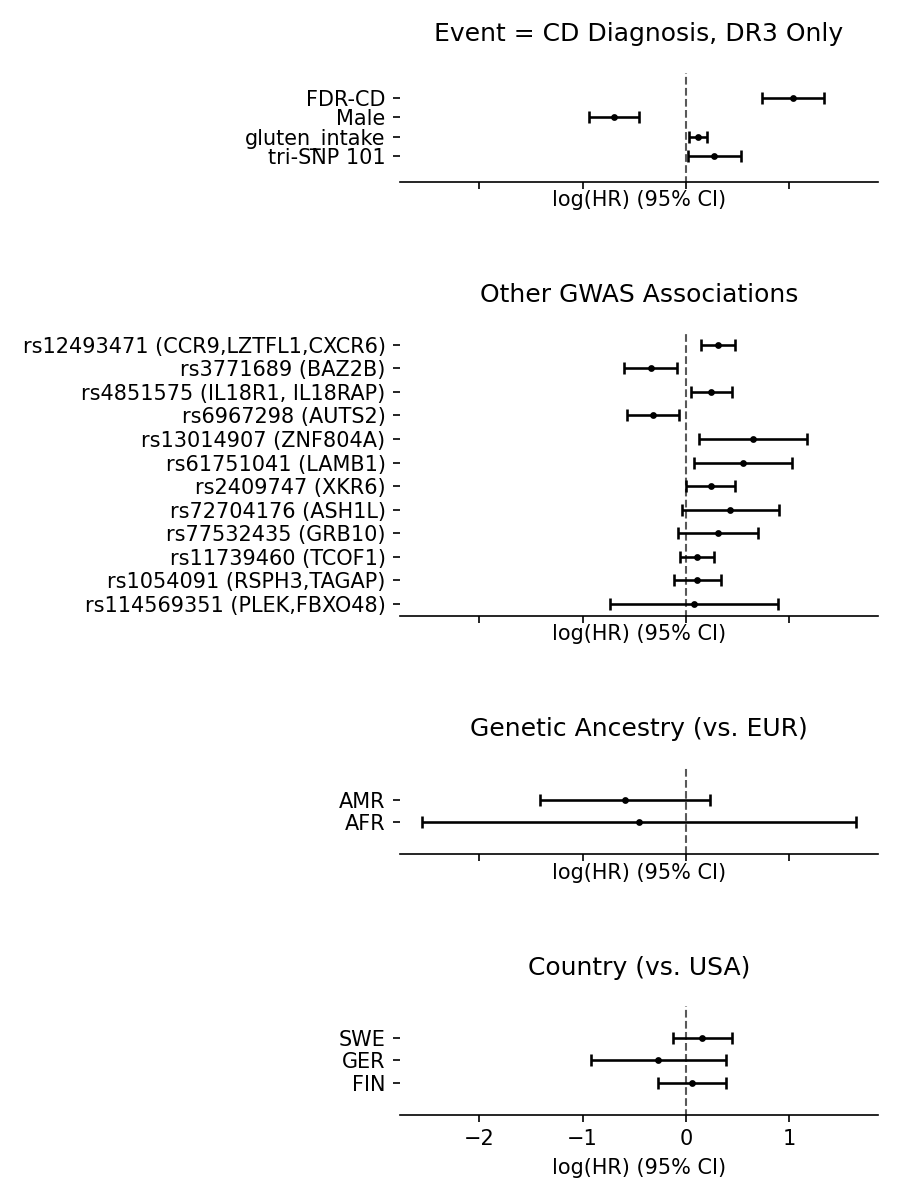

**Supplementary Figure 10.** Complete model output for CD outcome using only DR3-DQ2 homozygotes. log(HR) values and 95% CI are plotted for each covariate included in the model. Vertical line marks the log(HR)=0 (no change in risk). N_observations_= 1364, N_events_= 298.

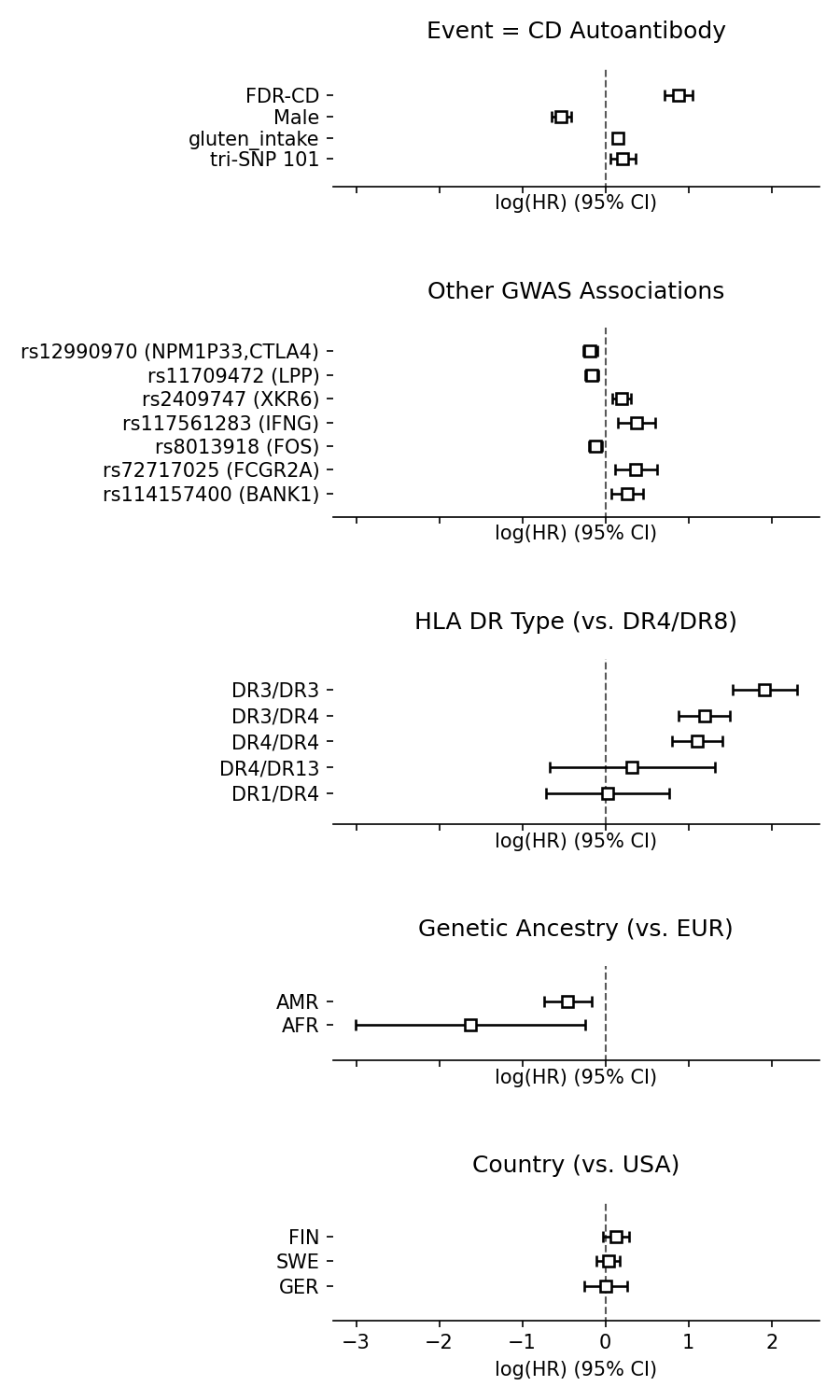

**Supplementary Figure 11.** Complete model output for CDA outcome. log(HR) values and 95% CI are plotted for each covariate included in the model. Vertical line marks the log(HR)=0 (no change in risk). N_observations_= 6557, N_events_= 1282.

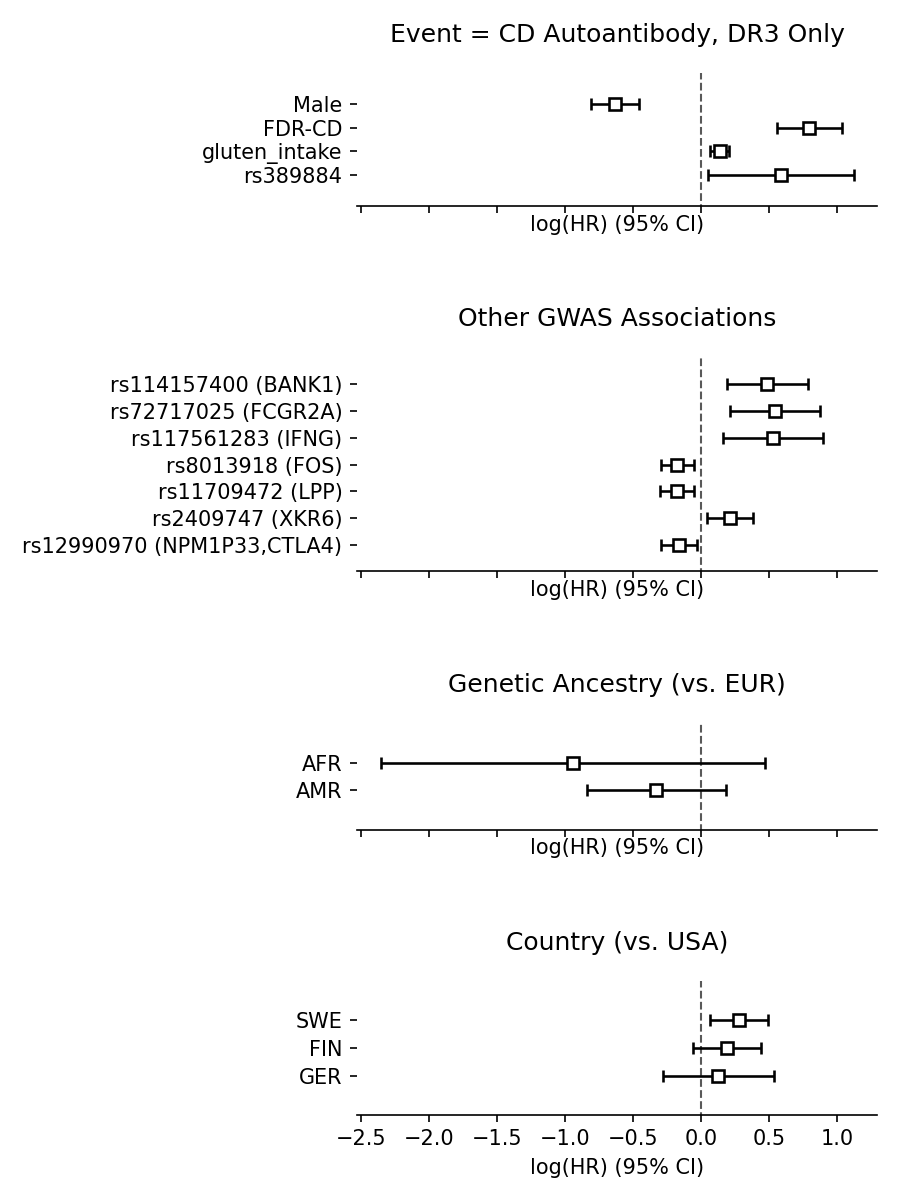

**Supplementary Figure 12.** Complete model output for CDA outcome using only DR3-DQ2 homozygotes. log(HR) values and 95% CI are plotted for each covariate included in the model. Vertical line marks the log(HR)=0 (no change in risk). N_observations_= 1370, N_events_= 526

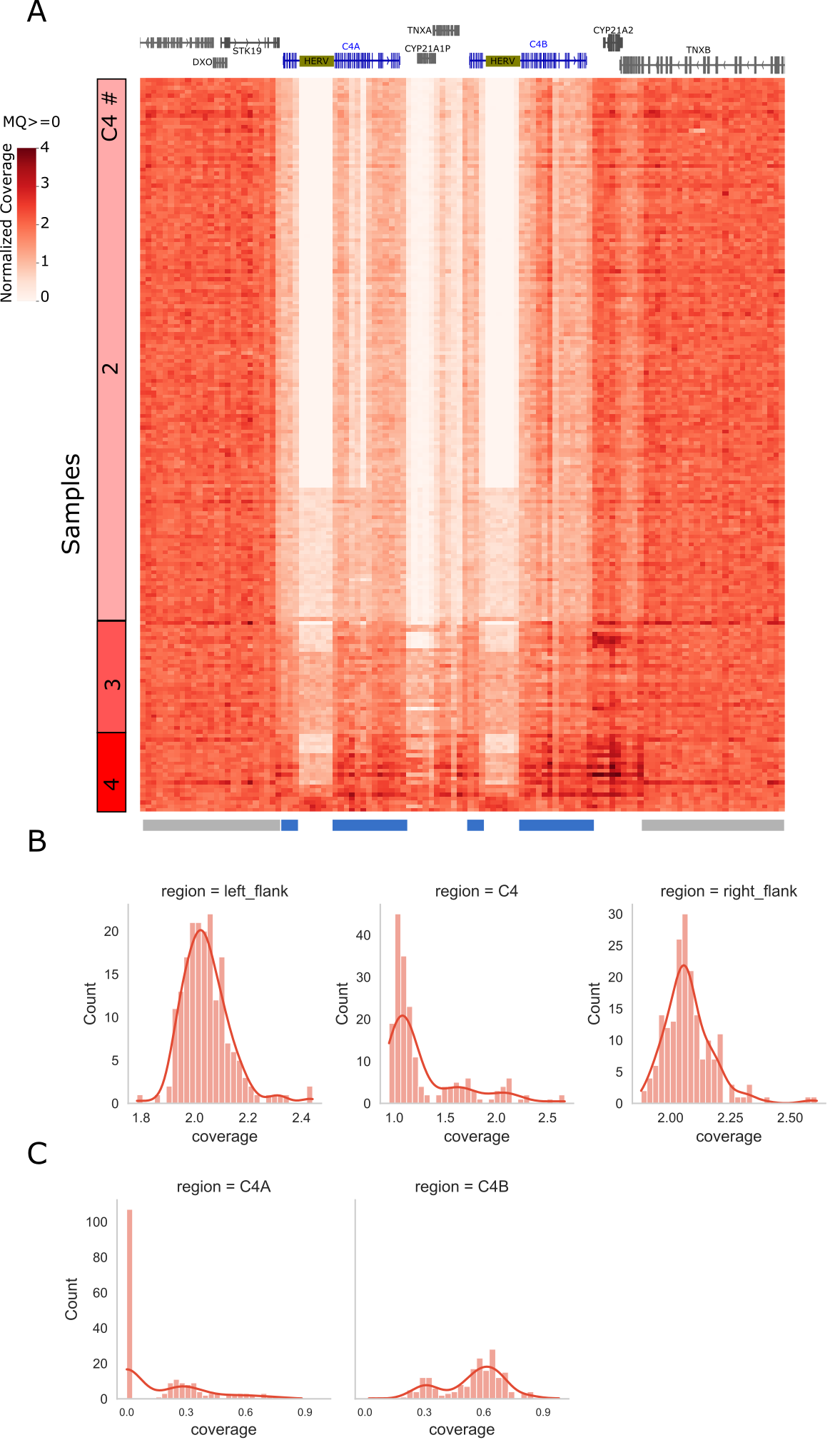

**Supplementary Figure 13. Sequence read coverage in C4 region.** (A) Normalized read coverage from WGS data of 188 homozygous DR3-DQ2 individuals. Reduced coverage of C4 genes relative to the flanking regions indicate the presence of frequent gene deletions. Total C4 copy numbers per sample indicated on the left side of the heatmap were estimated based on the average coverage per sample in C4 region demarcated with blue boxes below the heatmap, including both C4A and C4B but excluding reads mapping to intronic HERV insertion. A maximum value of 4 was used for the heatmap to moderate high outlier values. (B) Histograms and the kernel density estimates (kde) of the sample distribution based on average read coverage for C4 (blue boxes below heatmap) and copy number invariant flanking regions (gray boxes below heatmap). Flanking regions show a tight distribution with a peak around 2 indicating normal/diploid status. C4 coverage distribution shows 3 peaks around 1, 1.5 and 2; most likely corresponding to 2, 3 and 4 total copies. (C) Distribution of unique coverage corresponding to C4A and C4B genes (see Figure 3). Major C4A peak at 0 coverage indicating C4A null samples and peaks around 0.3 and 0.6 are likely 1 and 2 copies of C4A, respectively. C4B distribution shows a single null sample as well as 2 peaks around 0.3 and 0.6.
