## Supplementary material for "Polymorphisms in Intron 1 of HLA-DRA Differentially Associate with Type 1 Diabetes and Celiac Disease and Implicate Involvement of Complement System Genes C4A and C4B": TEDDY Study Group Appendix

### The TEDDY Study Group

**Colorado Clinical Center:** Marian Rewers, M.D., Ph.D., PI[1,4,6,9,10], Kimberly Bautista[11], Judith Baxter[8,911], Daniel Felipe-Morales, Brigitte I. Frohnert, M.D., Ph.D.[2,13], Marisa Stahl, M.D.[12], Isabel Flores Garcia, Patricia Gesualdo[2,6,11,13], Sierra Hays, Michelle Hoffman[11,12,13], Rachel Karban[11], Edwin Liu, M.D.[12], Leila Loaiza Jill Norris, Ph.D.[2,3,11], Holly O'Donnell, Ph.D.[8], Loana Thorndahl, Andrea Steck, M.D.[3,13], Kathleen Waugh[6,7,11]. University of Colorado, Anschutz Medical Campus, Barbara Davis Center for Childhood Diabetes, Aurora, CO, USA.

**Finland Clinical Center:** Jorma Toppari, M.D., Ph.D., PI[¥^1,4,10,13], Olli G. Simell, M.D., Ph.D., Annika Adamsson, Ph.D.^[11], Suvi Ahonen*±§, Mari Åkerlund*±§, Sirpa Anttila[µ¤], Leena Hakola*±, Anne Hekkala, M.D.[µ¤], Tiia Honkanen[µ¤], Heikki Hyöty, M.D., Ph.D.*±[6], Jorma Ilonen, M.D., Ph.D.¥[3], Sanna Jokipuu^, Taru Karjalainen[µ¤], Leena Karlsson^, Jukka Kero M.D., Ph.D.¥^[3, 13], Jaakko J. Koskenniemi M.D., Ph.D.¥^, Miia Kähönen[µ¤11,13], Mikael Knip, M.D., Ph.D.*±, Minna-Liisa Koivikko[µ¤], Katja Kokkonen*±, Merja Koskinen*±, Mirva Koreasalo*±§[2], Kalle Kurppa, M.D., Ph.D.*±[12], Salla Kuusela, M.D. [µ¤], Jarita Kytölä*±, Jutta Laiho, Ph.D.*[6], Tiina Latva-aho[µ¤], Siiri Leisku*±, Laura Leppänen^, Katri Lindfors, Ph.D.*[12], Maria Lönnrot, M.D., Ph.D.*±[6], Elina Mäntymäki^, Markus Mattila*±, Maija Miettinen§[2], Teija Mykkänen[µ¤], Tiina Niininen±*[11], Sari Niinistö§[2], Noora Nurminen*±, Sami Oikarinen, Ph.D.*±[6], Hanna-Leena Oinas*±, Paula Ollikainen[µ¤], Zhian Othmani¥, Sirpa Pohjola [µ¤], Solja Raja-Hanhela[µ¤], Jenna Rautanen±§, Anne Riikonen*±§[2], Minna Romo^, Juulia Rönkä[µ¤], Nelli Rönkä[µ¤], Satu Simell, M.D., Ph.D.¥[12], Päivi Tossavainen, M.D.[µ¤], Mari Vähä-Mäkilä¥, Eeva Varjonen^[11], Riitta Veijola, M.D., Ph.D.[µ¤13], Irene Viinikangas[µ¤], Silja Vilmi[µ¤], Suvi M. Virtanen, M.D., Ph.D.*±§[2]. ¥University of Turku, Turku, Finland, *Tampere University, Tampere, Finland, µUniversity of Oulu, Oulu, Finland, ^Turku University Hospital, Hospital District of Southwest Finland, Turku, Finland, ±Tampere University Hospital, Tampere, Finland, ¤Oulu University Hospital, Oulu, Finland, §Finnish Institute for Health and Welfare, Helsinki, Finland.

**Georgia/Florida Clinical Center:** Richard McIndoe, Ph.D., PI^[4,10], Desmond Schatz, M.D.*[4,7,8], Diane Hopkins^[11], Michael Haller, M.D.*[13], Risa Bernard^[11], Melissa Gardiner^[11], Ashok Sharma, Ph.D.^, Laura Jacobsen, M.D.*[13], Jennifer Hosford^, Kennedy Petty^, Leah Myers^, Chelsea Salmon*. ^Center for Biotechnology and Genomic Medicine, Augusta University, Augusta, GA, USA. *University of Florida, Pediatric Endocrinology, Gainesville, FL, USA.

**Germany Clinical Center:** Anette G. Ziegler, M.D., PI[1,3,4,10], Ezio Bonifacio Ph.D.*, Cigdem Gezginci, Willi Grätz, Anja Heublein, Eva Hohoff¥[2], Sandra Hummel, Ph.D.[2], Annette Knopff[7], Melanie Köger, Sibylle Koletzko, M.D.¶[12], Claudia Ramminger[11], Roswith Roth, Ph.D.[8], Jennifer Schmidt, Marlon Scholz, Joanna Stock[8,11,13], Katharina Warncke, M.D.[13], Lorena Wendel, Christiane Winkler, Ph.D.[2,11]. Forschergruppe Diabetes e.V. and Institute of Diabetes Research, Helmholtz Zentrum München, Forschergruppe Diabetes, and Klinikum rechts der Isar, Technische Universität München, Neuherberg, Germany. *Center for Regenerative Therapies, TU Dresden, Dresden, Germany, ¶Dr. von Hauner Children's Hospital, Department of Gastroenterology, Ludwig Maximillians University Munich, Munich, Germany, ¥University of Bonn, Department of Nutritional Epidemiology, Bonn, Germany.

**Sweden Clinical Center:** Åke Lernmark, Ph.D., PI[1,3,4,5,6,8,9,10], Daniel Agardh, M.D., Ph.D.[6,12], Carin Andrén Aronsson, Ph.D.[2,11,12], Rasmus Bennet, Corrado Cilio, Ph.D., M.D.[6], Susanne Dahlberg, Ulla Fält, Malin Goldman Tsubarah, Emelie Ericson-Hallström, Lina Fransson, Emina Halilovic, Gunilla Holmén, Susanne Hyberg, Berglind Jonsdottir, M.D., Ph.D.[11], Naghmeh Karimi, Helena Elding Larsson, M.D., Ph.D.[6,13], Marielle Lindström, Markus Lundgren, M.D., Ph.D.[13], Marlena Maziarz, Ph.D., Jessica Melin[11], Caroline Nilsson, Kobra Rahmati, Anita Ramelius, Falastin Salami, Ph.D., Anette Sjöberg, Evelyn Tekum Amboh Carina Törn, Ph.D.[3], Ulrika Ulvenhag, Terese Wiktorsson, Åsa Wimar[13]. Lund University, Lund, Sweden.

**Washington Clinical Center:** William A. Hagopian, M.D., Ph.D., PI[1,3,4,6,7,10,12,13], Michael Killian[6,7,11,12], Claire Cowen Crouch[11,13], Jennifer Skidmore[2], Trevor Bender, Megan Llewellyn, Cody McCall, Arlene Meyer, Jocelyn Meyer, Denise Mulenga[11], Nole Powell, Jared Radtke, Shreya Roy, Preston Tucker. Pacific Northwest Research Institute, Seattle, WA, USA.

**Pennsylvania Satellite Center:** Dorothy Becker, M.D., Margaret Franciscus, MaryEllen Dalmagro-Elias Smith[2], Ashi Daftary, M.D., Mary Beth Klein, Chrystal Yates. Children's Hospital of Pittsburgh of UPMC, Pittsburgh, PA, USA.

**Data Coordinating Center:** Jeffrey P. Krischer, Ph.D., PI[1,4,5,9,10], Rajesh Adusumali, Sarah Austin-Gonzalez, Maryouri Avendano, Sandra Baethke, Brant Burkhardt, Ph.D.[6], Martha Butterworth[2], Nicholas Cadigan, Joanna Clasen, Kevin Counts, Laura Gandolfo, Jennifer Garmeson, Veena Gowda, Christina Karges, Shu Liu, Xiang Liu, Ph.D.[2,3,8,13], Kristian Lynch, Ph.D. [6,8], Jamie Malloy, Lazarus Mramba, Ph.D.[2], Cristina McCarthy[11], Jose Moreno, Hemang M. Parikh, Ph.D.[3,8], Cassandra Remedios, Chris Shaffer, Susan Smith[11], Noah Sulman, Ph.D., Roy Tamura, Ph.D.[1,2,11,12,13], Dena Tewey, Henri Thuma, Michael Toth, Ulla Uusitalo, Ph.D.[2], Kendra Vehik, Ph.D.[4,5,6,8,13], Ponni Vijayakandipan, Melissa Wroble, Jimin Yang, Ph.D., R.D.[2], Kenneth Young, Ph.D. *Past staff: Michael Abbondondolo, Lori Ballard, Rasheedah Brown, David Cuthbertson, Stephen Dankyi, Christopher Eberhard, Steven Fiske, David Hadley, Ph.D., Kathleen Heyman, Belinda Hsiao, Francisco Perez Laras, Hye-Seung Lee, Ph.D., Qian Li, Ph.D., Colleen Maguire, Wendy McLeod, Aubrie Merrell, Steven Meulemans, Ryan Quigley, Laura Smith, Ph.D.* University of South Florida, Tampa, FL, USA.

**Project scientist:** Beena Akolkar, Ph.D.[1,3,4,5,6,7,9,10]. National Institutes of Diabetes and Digestive and Kidney Diseases, Bethesda, MD, USA.

**Autoantibody Reference Laboratories:** Liping Yu, M.D.^[5], Dongmei Miao, M.D.^, Kathleen Gillespie*[5], Kyla Chandler*, Ilana Kelland*, Yassin Ben Khoud*, Matthew Randell *. ^Barbara Davis Center for Childhood Diabetes, University of Colorado Denver, *Bristol Medical School, University of Bristol, UK.

**Genetics Laboratory:** Stephen S. Rich, Ph.D.[3], Wei-Min Chen, Ph.D.[3], Suna Onengut-Gumuscu, Ph.D.[3], Emily Farber, Rebecca Roche Pickin, Ph.D., Jonathan Davis, Jordan Davis, Dan Gallo, Jessica Bonnie, Paul Campolieto. Center for Public Health Genomics, University of Virginia, Charlottesville, VA, USA.

**HLA Reference Laboratory:** William Hagopian[3], M.D., Ph.D., Jared Radtke, Preston Tucker. Pacific Northwest Research Institute, Seattle, WA, USA. (Previously Henry Erlich, Ph.D.[3], Steven J. Mack, Ph.D., Anna Lisa Fear. Center for Genetics, Children's Hospital Oakland Research Institute.)

**Repository:** Chris Deigan. NIDDK Biosample Repository at Fisher BioServices, Rockville, MD, USA. (Previously Ricky Schrock, Polina Malone, Sandra Ke, Niveen Mulholland, Ph.D.)

**Other contributors:** Thomas Briese, Ph.D.[6], Columbia University. Todd Brusko, Ph.D.[5], University of Florida, Gainesville, FL, USA. Teresa Buckner, Ph.D.[2], University of Northern Colorado, Greeley, CO. Suzanne Bennett Johnson, Ph.D.[8,11], Florida State University, Tallahassee, FL, USA. Eoin McKinney, Ph.D.[5], University of Cambridge, Cambridge, UK. Tomi Pastinen, M.D., Ph.D.[5], The Children's Mercy Hospital, Kansas City, MO, USA. Steffen Ullitz Thorsen, M.D., Ph.D.[2], Department of Clinical Immunology, University of Copenhagen, Copenhagen, Denmark, and Department of Pediatrics and Adolescents, Copenhagen University Hospital, Herlev, Denmark. Eric Triplett, Ph.D.[6], University of Florida, Gainesville, FL, USA.

***Committees:***
[1]Ancillary Studies, [2]Diet, [3]Genetics, [4]Human Subjects/Publicity/Publications, [5]Immune Markers, [6]Infectious Agents, [7]Laboratory Implementation, [8]Psychosocial, [9]Quality Assurance, [10]Steering, [11]Study Coordinators, [12]Celiac Disease, [13]Clinical Implementation.
